# A latent outcome variable approach for Mendelian randomization using the expectation maximization algorithm

**DOI:** 10.1101/2024.08.24.24312485

**Authors:** Lamessa Dube Amente, Natalie T Mills, Thuc Duy Le, Elina Hyppönen, S Hong Lee

**Affiliations:** Australian Centre for Precision Health, University of South Australia, Adelaide, SA 5000, Australia; UniSA Allied Health and Human Performance, University of South Australia, Adelaide, SA 5000, Australia; South Australian Health and Medical Research Institute, Adelaide, SA 5000, Australia; Epidemiology department, Jimma University, Jimma, 378, Ethiopia; Discipline of Psychiatry, University of Adelaide, Adelaide, South Australia 5000, Australia; UniSA STEM, University of South Australia, Mawson Lakes, SA 5095, Australia; UniSA Clinical and Health Sciences, University of South Australia, Adelaide, SA 5000, Australia

## Abstract

Mendelian randomization (MR) is a widely used tool to uncover causal relationships between exposures and outcomes. However, existing MR methods can suffer from inflated type I error rates and biased causal effects in the presence of invalid instruments. Our proposed method enhances MR analysis by augmenting latent phenotypes of the outcome, explicitly disentangling horizontal and vertical pleiotropy effects. This allows for explicit assessment of the exclusion restriction assumption and iteratively refines causal estimates through the expectation-maximization algorithm. This approach offers a unique and potentially more precise framework compared to existing MR methods. We rigorously evaluate our method against established MR approaches across diverse simulation scenarios, including balanced and directional pleiotropy, as well as violations of the Instrument Strength Independent of Direct Effect (InSIDE) assumption. Our findings consistently demonstrate superior performance of our method in terms of controlling type I error rates, bias, and robustness to genetic confounding. Additionally, our method facilitates testing for directional horizontal pleiotropy and outperforms MR-Egger in this regard, while also effectively testing for violations of the InSIDE assumption. We apply our method to real data, demonstrating its effectiveness compared to traditional MR methods. This analysis reveals the causal effects of body mass index (BMI) on metabolic syndrome (MetS) and a composite MetS score calculated by the weighted sum of its component factors. While the causal relationship is consistent across most methods, our proposed method shows fewer violations of the exclusion restriction assumption, especially for MetS scores where horizontal pleiotropy persists and other methods suffer from inflation.

## Introduction

Complex traits result from the interplay of genetic and environmental factors^(1-3)^. Genome-wide association studies (GWAS) have provided compelling evidence for the genetic underpinnings of these traits, while epidemiological studies have underscored the role of environmental factors. Often, these factors intersect, influencing traits through both horizontal and vertical pleiotropy^(4)^.

Horizontal pleiotropy, also known as “direct pleiotropy” or simply “pleiotropy,” occurs when a single genetic variant influences multiple traits. In contrast, vertical pleiotropy, termed “indirect pleiotropy” or “mediated pleiotropy,” emerges when a genetic variant is linked to variables sharing the same causal pathway to the outcome^(5)^. Like horizontal pleiotropy, which affects variables on different causal pathways leading to the outcome, vertical pleiotropy holds particular significance in epidemiological and clinical contexts for several reasons^(4, 6, 7)^:

1. Vertical pleiotropy plays a pivotal role in disease susceptibility, where genes affecting immune function can indirectly influence the risk of various infectious or autoimmune diseases.
2. Understanding vertical pleiotropy is crucial for predicting and optimizing treatment responses, as genes influencing drug metabolism can significantly impact individual responses to specific medications.
3. Knowledge of vertical pleiotropy can enhance diagnostic accuracy by identifying shared genetic factors underlying multiple related phenotypes, leading to more effective screening and diagnostic strategies for complex diseases or conditions.
4. Vertical pleiotropy supports pathway-based approaches in epidemiology and clinical research^(3)^, wherein genetic variants affecting multiple traits within the same biological pathway or process are collectively analysed. This approach enhances our understanding of disease mechanisms and informs the development of targeted interventions.

Mendelian randomization (MR) stands as a valuable tool for detecting vertical pleiotropy and estimating the causal effects of exposure on the outcome^(8-11)^. In MR, careful selection of vertical pleiotropy variants as instrumental variables (IVs) is essential. Valid IVs must hold several criteria: they associate with the risk factor under investigation (relevance assumption), have no common cause with the outcome (independence assumption), and solely influence the outcome through the risk factor (exclusion restriction assumption)^(3)^. Several methodologies have been developed to address these challenges in MR analysis, including MR-Egger regression^(12)^, weighted mode MR^(13)^, weighted median MR^(14)^, MR Pleiotropy RESidual Sum and Outlier (MR-PRESSO)^(15)^, Generalized Summary-data-based Mendelian Randomization (GSMR)^(16)^, MR using mixture models (MR-Mix)^(17)^, contamination mixture (MR-ConMix)^(11)^ and Iterative Mendelian Randomization and Pleiotropy (IMRP) approach ^(18)^.

Among these methods, advanced methods such as MR-Mix, MR-ConMix and IMRP are noteworthy for the ability to simultaneously conduct MR analysis and identify horizontal pleiotropy variants as invalid instruments based on GWAS summary statistics^(11, 18)^. IMRP models the residual distribution of 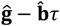, where **ĝ** and 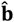 represent the effect sizes of an IV for an outcome and an exposure, respectively, and *τ* is the causal effect of the exposure on the outcome. By identifying pleiotropic IVs and recalculating the causal effect after excluding them at each iterative step, the method enables simultaneous MR analysis and detection of pleiotropic variants. The MR-Mix and MR-ConMix methods construct a likelihood function accounting for different distributions of valid and invalid IVs. The likelihood is maximized over various causal effect values and configurations, minimising 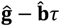. Importantly, these methods have demonstrated superior performance compared to existing MR methods in extensive simulations ^(18-21)^.

While our proposed method shares similarities with existing approaches, it also has distinct differences. Unlike other methods, which assume that the residual follows a mixture of normal distributions depending on the valid or invalid status of IVs, our proposed method generates latent phenotypes of the outcome that explicitly exclude vertical pleiotropy effects and iteratively refines *τ* through the expectation and maximisation (EM) algorithm. Hence, the proposed method provides a unique and potentially more precise framework for MR analysis compared to conventional methods. Additionally, we provide explicit theoretical derivations demonstrating that the proposed method can be applied using summary statistics. We compare our proposed method with existing MR methods and demonstrate that it performs better when the InSIDE assumption is violated, and a significant proportion of the IVs are invalid. Under other conditions, its performance is comparable to that of existing methods. Finally, we introduce statistical tests for assessing directional pleiotropy and InSIDE assumption violation, which can help in quantifying the validity of the analysis.

## Materials and methods

### MR models

MR is a statistical method used to assess causal relationships between an exposure (such as a modifiable risk factor or treatment) and an outcome (such as a disease or health outcome) using genetic variants as IVs. The basic principle of MR relies on the random allocation of genetic variants at conception, which is analogous to the randomization process in a randomized controlled trial^(3)^. This random allocation ensures that genetic variants are not influenced by confounding factors and allows for the estimation of causal effects.

The MR model typically involves three key components: 1) Genetic instruments: These are genetic variants that are associated with the exposure of interest (e.g., blood pressure, cholesterol levels) and are assumed to influence the outcome only through their effect on the exposure, i.e. excluding any horizontal pleiotropy variants. Valid genetic instruments should satisfy certain assumptions, including relevance (association with the exposure), independence from confounders, and absence of direct effects on the outcome other than through the exposure. 2) Exposure: This refers to the risk factor or treatment under investigation, which is associated with and instrumented by the genetic variants used in the MR analysis. 3) Outcome: This refers to the health outcome or disease of interest, which may be causally influenced by the exposure.

The causal model can be written as

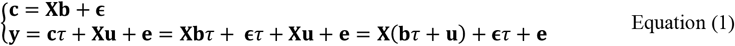

where **y** and **c** represent outcome and exposure traits, respectively, *τ* represents fixed causal effects of **c** on **y, X** is the genome-wide SNP genotype matrix, **b** and **u** are SNP effects associated with **c** and **y**, respectively, and **ϵ** and **e** encompass non-genetic residual effects pertaining to **c** and **y**, respectively. For simplicity, we denote **g = b***τ* + **u**, representing the overall genetic effects of **y**.

It is important to note that the genetic covariance between the exposure variable (**c**) and the outcome variable (**y**) is influenced by various factors. Specifically, the covariance between the genetic effects associated with the exposure (**b**) and the genetic effects associated with the outcome (**g = b***τ* + **u**) can be decomposed into two components: *cov*(**b, g**) **=** *var*(**b**)*τ* + *cov*(**b, u**) where *var*(**b**)*τ* arises from vertical pleiotropy, and *cov*(**b, u**) represents horizontal pleiotropy.

In situations where there is no horizontal pleiotropy (i.e., using valid IVs only), the value of *τ* can be estimated as *τ* **=** *cov*(**b, g**)*/var*(**b**). This estimation can be achieved using methods such as two-stage least squares (2SLS) or inverse variance weighting (IVW), especially when weights (e.g., standard errors) are available^(2)^. Additionally, advanced methods such as weighted median^(14)^, MR-Egger regression^(12)^, MR-PRESSO^(15)^, GSMR^(16)^, MR-Mix^(17)^, contamination mixture^(11)^ and IMRP^(18)^ can be employed in MR analyses. As previously reviewed^(18, 19, 21)^, there are cons and pros among the methods.

MR-Mix^(17)^ employs a novel approach of handling horizontal pleiotropy through a mixture modelling framework. Here, IVs are categorized into distinct subpopulations, each characterized by unique causal effects on the outcome, including their effect sizes and pleiotropic impacts. MR-Mix effectively estimates the proportion of IVs within each subpopulation along with their corresponding causal effects, facilitated by the use of maximum likelihood estimation to minimize the function 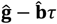 where **ĝ** and 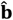 represent the estimated effect sizes of an IV for an outcome and an exposure, respectively, and *τ* is the causal effect of the exposure on the outcome. MR-Mix typically performs a two-step procedure. First, they fit a mixture model to the summary statistics obtained from GWAS data, estimating the parameters of the subpopulations and their causal effects. Second, they use these estimates to perform MR analysis, estimating the causal effect of the exposure on the outcome while accounting for horizontal pleiotropy.

Similarly, the contamination mixture model^(11)^ assumes that genetic variants can be classified into two distinct subpopulations: a “contaminated” subpopulation, where variants exhibit horizontal pleiotropy and influence the outcome directly, and a “valid instrument” subpopulation, where variants only affect the outcome through their association with the exposure. A likelihood function is constructed from the model, where valid IVs are assumed to have a normal distribution around the true causal effect, *τ*, while invalid IVs are assumed to have a normal distribution around zero. The likelihood is maximized over different values of the causal effect and configurations of valid and invalid IVs. If the assumption is satisfied, then the two sets of regression coefficients will satisfy a proportional relationship in the form 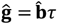. The causal estimate that maximizes the profile likelihood is then determined.

IMRP^(18)^ is an iterative method to simultaneously conduct MR analysis and identify horizontal pleiotropy variants as invalid instruments based on GWAS summary statistics. In each iteration, IMRP applies a t-test, 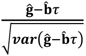, for each IV to determine pleiotropic IVs. By identifying pleiotropic IVs and recalculating the causal effect after excluding them at each iterative step, IMRP enables simultaneous MR analysis and detection of pleiotropic variants.

These methods offer several advantages over traditional MR methods by explicitly addressing horizontal pleiotropy, leading to more accurate estimates of causal effects, especially in the presence of pleiotropic genetic variants.

### Latent Outcome Variable Approach (LOVA) for MR using the Expectation Maximization (EM) steps

Suppose we know the true *τ* without error. According to equation (1), a new set of phenotypes can be generated as follows:

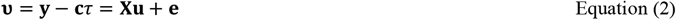

The new phenotypes, denoted by **υ**, exclude the vertical pleiotropy effects (**c***τ*), allowing only for horizontal pleiotropy effects with residual effects. By using **υ**, the GWAS summary statistics will distinctly reveal which variants are not associated with the outcome, satisfying the exclusion restriction assumption, a key aspect in MR.

However, given the true *τ* is unknown, we propose employing an iterative EM algorithm:

1. **Initialization**: Start with initial estimates for *τ*^(*t*)^ where *t* is the iteration number (*t***=**0, 1, 2, …)
2. **Expectation (E) Step**: Calculate the expected values of the latent variable **υ**^(*t*+1)^ **= y** − **c***τ*^(*t*)^ **= y** − (**Xb** + **ϵ**)*τ*^(*t*)^ given the observed data and the current parameter estimates. This step involves imputing the latent values based on the observed data and the current estimates of *τ*. The calculation of **υ** should incorporate both the observed data **y** and the linear predictor (**Xb** + **ϵ**)*τ*^(*t*)^, representing the expected value of **υ** given the current estimate of *τ*.
3. **Maximization (M) Step**: Update the parameter estimates, *τ*, to maximize the expected log-likelihood computed in the E-step, i.e. log-L(*τ* | **y, υ, I**) where I is the inclusion indicator for vertical pleiotropy (0 or 1). This step first involves fitting a regression model to estimate the parameter, *û*_*j*_, using the imputed **υ** values. Then, *τ* can be estimated using an IVW model, represented as 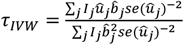 where Ij is the inclusion indicator with I_j_=1 if the significance criteria are met; otherwise I_j_=0 (see below).
4. **Iteration**: Repeat steps 2 and 3 until convergence, where the parameter estimates no longer change significantly between iterations or a convergence criterion is met.

In the EM steps, two key parameters need to be specified by the users: the initial value for *τ* and the significance level used to select vertical pleiotropy variants (referred to as the inclusion indicator) in each iteration.

### Initial value for *τ* in the EM method

In many scenarios, it is often considered reasonable to start with the assumption of no causal effect and initialize the parameter *τ* to 0 in the EM method. This initial value implies that there is no vertical pleiotropy variant influencing the outcome. By beginning with this assumption, the analysis starts from a neutral standpoint, which can help prevent the algorithm from being biased towards finding spurious causal relationships. Consequently, initializing *τ* to 0 can contribute to reducing the likelihood of encountering false positives, where associations are erroneously detected between variables that do not have a genuine causal relationship.

However, it is important to acknowledge that prior knowledge about the presence or absence of vertical pleiotropy variants and the true nature of *τ* can greatly inform the choice of initial values. If reliable information exists regarding specific variants that may exert vertical pleiotropy effects or if there are well-established estimates for *τ* based on previous research or biological understanding, it may be advantageous to use these values as the initial starting point for the EM algorithm. Incorporating such prior knowledge can potentially enhance the accuracy of the estimation process by guiding the algorithm towards more relevant regions of the parameter space.

### Significant criteria for the inclusion indicator in the M step

The significance level chosen for the inclusion indicator serves as a critical determinant in the process. It sets the threshold for identifying vertical pleiotropy variants. This involves carefully assessing the statistical significance of each variant’s effect on both the outcome and the exposure of interest.

To implement this criterion, variants exhibiting significant effects on the latent outcome variable (i.e., outcome corrected for vertical pleiotropy based on eq. 2) are typically excluded from the analysis, as they may introduce bias by confounding the relationship between the exposure and outcome (exclusion restriction assumption). Similarly, variants with non-significant effects on the exposure are also excluded, as they are less likely to contribute to any genuine mediation effects (relevance assumption).

By applying these criteria, researchers can effectively filter out irrelevant genetic variants and focus only on variants that have the potential to influence the causal pathway under investigation. This approach enhances the validity and reliability of the MR analysis, ensuring that the identified genetic instruments are robust and capable of providing meaningful insights into causal relationships. As a default, we employ the following criteria: if the estimated parameter *û*_*j*_ has a p-value > 0.05 and 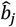 has a p-value < 5E-08 then I_j_=1; otherwise, I_j_=0.

### MR-LOVA based on GWAS summary statistics

In the E step for each iteration, the calculation of **υ** involves the following equation:

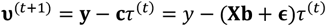

This calculation necessitates individual-level data, as also shown in eq. (1) (i.e. **y, c, X**). However, the proposed method can use GWAS summary stats solely, not relying on individual level data.

Without loss of generality, assuming that SNP genotypes are column-standardized and SNP effects estimated from the outcome and exposure GWAS are standardized, the updated SNP effects based on the corrected phenotypes, **υ**, can be derived as

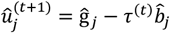

and, the sampling variance of the updated SNP effects for the outcome can be expressed as

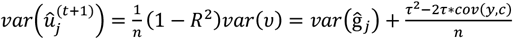

where R^2^ is typically close to zero, owing to a single genetic variant, *var*(*v*) **=** *var*(*y*) + *var*(*c*) ∗ *τ*^2^ − 2*τ* ∗ *cov*(*y, c*) **=** 1 + *τ*^2^ − 2*τ* ∗ *cov*(*y, c*), and 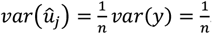. Note that *var*(ĝ_*j*_) is assumed to be known from GWAS summary statistics, and *cov(y,c)* can be approximated as *cov* 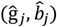,assuming that correlations among genetic variants are negligible, i.e. using approximately independent SNPs after pruning or clumping^(22)^. Therefore, given updated 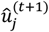 and *var*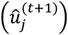, the EM algorithm can be operated.

The assumption of negligible correlations among genetic variants can be relaxed by incorporating the correlation structure among SNPs from a reference panel data (1KG or HapMap). Following^(23)^, the estimated regression coefficients from a multiple regression that accounts for the correlation between explanatory variables are given by 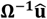 where **Ω** is the correlation matrix among IVs. The variance of 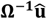 is expressed as:

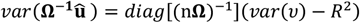

where *var*(*v*) **=** 1 + *τ*^2^ − 2*τ* ∗ *cov*(*y, c*) and 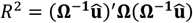, which is not negligible due to the presence of multiple genetic variants.

### Permutation

Permutation testing is often paired with the EM algorithm, particularly in scenarios where conventional statistical assumptions may not apply or when the null distribution of a test statistic is unknown. This method involves iteratively reshuffling the labels of observed data points to create a null distribution for the test statistic. By comparing the observed test statistic to the distribution derived from these permutations, one can gauge the significance of the observed result. Permutation testing offers a non-parametric approach to hypothesis testing, enabling control over the type I error rate under the null hypothesis without necessitating specific distributional assumptions. In our approach, we employ permutation by shuffling both the estimated parameter *û*_*j*_ and its standard error.

### Statistical test for directional pleiotropy and InSIDE assumption violation

The proposed method, which estimates direct SNP effects (**u** in eq. (1)) based on latent outcome variables (**υ**), facilitates tests for both directional pleiotropy and the InSIDE assumption violation.

The directional pleiotropy test is performed using a one-sample t-test to determine if the mean of **u** is significantly different from zero. A significant p-value indicates directional pleiotropy, while a non-significant p-value suggests its absence. This test is compared with the MR-Egger method, a standard approach for detecting directional pleiotropy.

The InSIDE assumption violation test assesses the correlation between direct SNP effects on the outcome and genetic effects on the exposure, i.e. cor(**u**,**b**). A Pearson correlation test determines if cor(**u**,**b**) significantly differs from zero. A significant p-value indicates a violation of the InSIDE assumption, whereas a non-significant p-value suggests no violation.

### Simulation

To demonstrate the performance of the proposed method compared with existing MR methods, we conducted a simulation study. Following previous studies^(19, 24)^, we used four distinct scenarios, 1) no pleiotropy, 2) balanced pleiotropy, 3) directional pleiotropy and 4) pleiotropy through a confounder. The Instrument Strength Independent of Direct Effect (InSIDE) assumption holds true for all scenarios except the fourth.

We simulated data according to Eq (1), with the additional consideration of a confounder as follows:

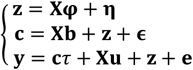

where **z** is a confounder that is often unobserved, **φ** represents SNP effects associated with **z, η** denotes non-genetic residual effects pertaining to **z**, and other terms are already defined as above.

Following^(19, 24)^, the entries of the *n*x*m* genotype matrix **X** were independently drawn from a Binomial distribution B(2, *f*_*i*_). For each SNP *i*, its MAF *f*_*i*_ was determined independently from others using a uniform distribution U(0.1, 0.3).

The IV strengths **b** were generated from a left-truncated normal distribution. For *m* = 30, IV strengths were generated from *N*(0, 0.1) left-truncated at 0.1. For *m* = 100, they were from N(0, 0.05) left-truncated at 0.05.

When considering scenarios involving pleiotropy (2 – 4), the details are as follows:

Scenario 2: Balanced pleiotropy and InSIDE satisfied, where pleiotropy effects **u** were generated independently from N(0,0.15) and **φ** = 0;

Scenario 3: Directional pleiotropy and InSIDE satisfied, where **u** were generated from N(0.1, 0.075) and **φ** = 0;

Scenario 4: Directional pleiotropy and InSIDE violated, where **u** were generated from N(0.1, 0.075) and **φ**were generated from U(0; θ)

**η, ϵ, e** were generated from N(0, 1) independently.

The summary data for genetic associations were calculated for the exposure and the outcome on non-overlapping sets of individuals, each consisting of n individuals. For scenarios (2) and (3), *τ* was varied between 0 and an expected value of 0.2 when both the outcome and exposure are scaled (i.e., the true *τ* under unscaled traits is *τ* scaled multiplied by the standard error of the outcome divided by the standard error of the exposure). The investigation included different proportions of invalid IVs: 0, 0.3, 0.5, and 0.7, with sample sizes for exposure and outcome set at 10,000 or 50,000. For scenario (4), We investigated various levels of correlated pleiotropy effects by varying θ across three values: 0.1, 0.4, and 0.7.

### Real data analysis

We applied the proposed method (MR-LOVA) to analyse publicly available GWAS summary-level data to explore causal relationships underlying variety of exposure-outcome pairs of interest. We assessed data for coronary heart disease CAD)^(25)^ as the outcome and analysed its relationship with blood lipids^(26)^, blood pressure^(27)^ as exposures. We selected these pairs based on sample sizes, number of instruments, and existing evidence from epidemiological and recent MR studies^(17, 18, 20, 24, 28)^.

In addition, we analysed the relationship between BMI^(29)^ as an exposure and MetS^(30, 31)^ as an outcome. Although obesity and MetS are genetically correlated^(31, 32)^, they are distinct phenotypes. For instance, some individuals are metabolically healthy despite being obese, while others are metabolically unhealthy despite having a normal weight^(33)^. This distinction makes it particularly interesting to explore whether obesity directly causes MetS^(34-36)^. Pairs with high genetic correlations, such as BMI and MetS, serve as a valuable case for comparing methods, as they are prone to horizontal pleiotropy, which mirrors the simulation scenario 4, where the InSIDE assumption is violated. While the causal relationship between obesity and MetS is an important question, our primary focus here is on comparing methodological approaches

For MetS, we extracted GWAS summary data for both binary and continuous outcomes. The binary outcome is referred to as MetS, and the continuous outcome is termed MetS score. MetS (binary) was defined based on the International Diabetes Federation criteria^(37)^, with individuals classified as having MetS if they met three or more of the following criteria: central obesity (WC ≥ 88 cm in females and ≥ 102 cm in males), elevated fasting triglycerides (TG) levels (≥ 1.7 mmol/L or medication for elevated TG), reduced high density lipoprotein cholesterol (HDL-C) (< 1.29 mmol/L in females and < 1.03 mmol/L in males or medication for reduced HDL-C), elevated blood pressure (systolic blood pressure/diastolic blood pressure (SBP/DBP) ≥ 130/85 mmHg), and elevated fasting glucose (≥ 5.6 mmol/L or medication for elevated glucose). The MetS score, a weighted sum of scores from the five components based on the genomic structural equation modelling^(38)^, captures genetic correlations and shared genetic variance among the components^(31)^. MetS scores provide potentially richer information than the binary MetS classification^(31, 39)^. GWAS summary data for MetS score was obtained from the Complex Trait Genetics Lab (CTG) (https://ctg.cncr.nl/software/summary_statistics), which contains 461,920 valid subjects of European ancestry.

Genetic correlation between the traits were estimated using LD regression score^(40)^. For MR analysis, we only selected SNPs as potential instruments if they reached genome-wide significance (p-value < 5 × 10^−8^) in the exposure GWAS. Furthermore, we used LD clumping with an r^2^ threshold of 0.1 and a window size of 500kb to select a set of independent instruments for each trait.

## Results

### Simulation results

Under the null hypothesis (no causal effects) and using valid instruments only (simulation scenario 1), all methods demonstrate well-controlled type I error rates, minimal mean squared error (MSE), and mean estimated causal effects close to zero (Figure 1).

**Figure 1:**
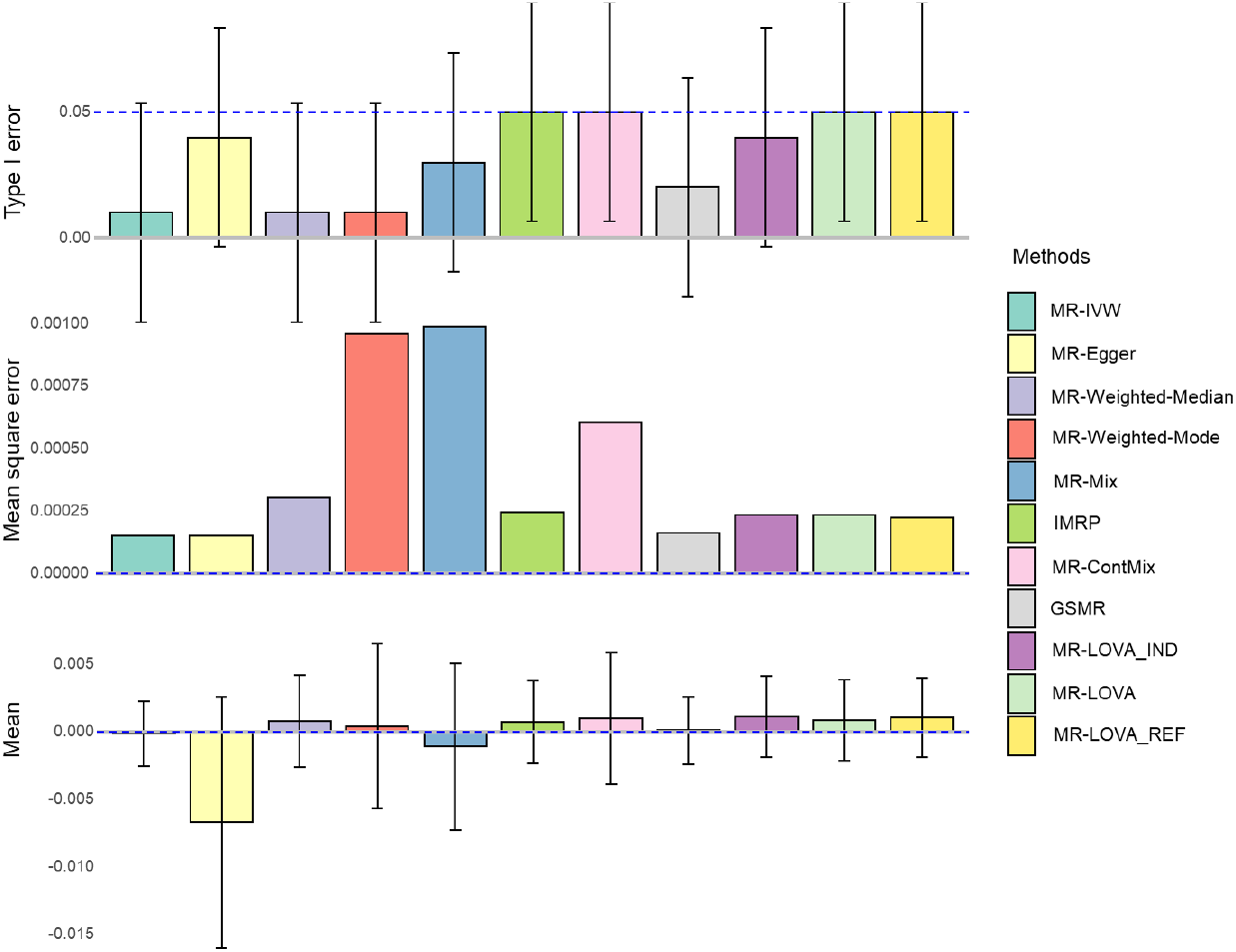
Simulation results under the null hypothesis with all valid instrumental variants. The blue dashed lines represent expected values, and error bars indicate 95% confidence intervals derived from 100 replicates. The sample size is 50,000. The number of SNPs is 100, all of which were initially provided to each method. Abbreviations: MR-IVW inverse variance weighted, MR-ConMix contamination mixture, GRSM generalized summary data based Mendelian randomization, IMRP iterative Mendelian randomization and pleiotropy, MR-LOVA_IND Mendelian randomization latent outcome variable approach based on individual-level data, MR-LOVA is based on summary-level data assuming independent variants, MR-LOVA_REF uses genotype reference panel to account for correlations between variants.

In simulation scenario 2 (balanced pleiotropy), an increase in the proportion of invalid instruments leads to noticeable inflation in overall type I error rates (Figure 2A) and an increase in MSE (Figure 2B), while maintaining unbiased mean estimates of causal effects (Figure 3C). Notably, MR methods like weighted median, weighted mode, MR-PRESSO, IMRP and GSMR exhibit substantial inflation in type I error rates when the proportion of invalid variants increase (Figure 2A), whereas MR-Egger and IVW show larger MSE (Figure 2B). Conversely, our proposed method (MR-LOVA) demonstrates minimal inflation of type I error rates and MSE, even when the proportion of invalid instruments reaches 70% (Figure 2A&B). Additionally, our method includes a permutation test capability to ensure control of type I error rates in most cases (Supplementary Figure 1).

**Figure 2:**
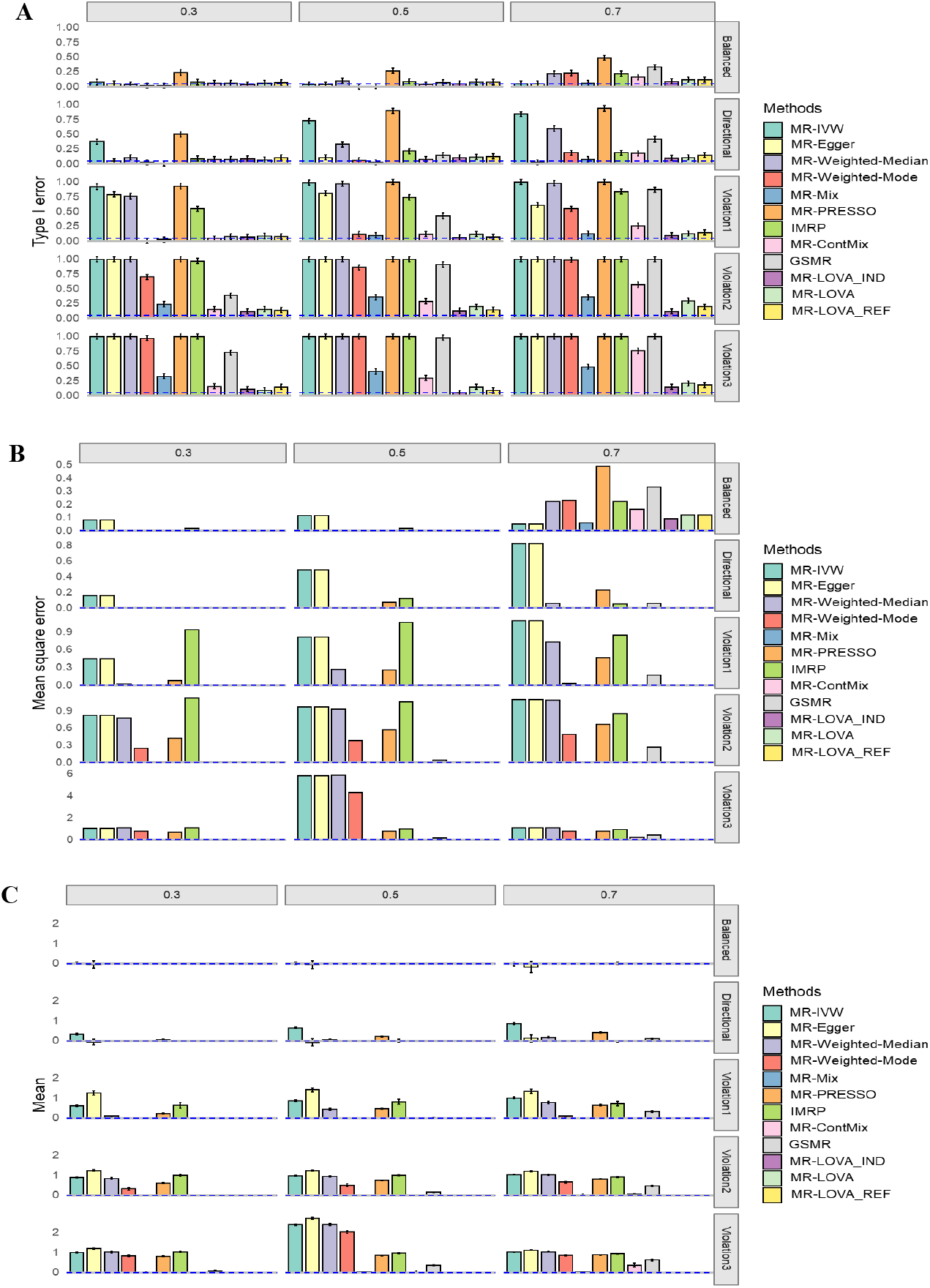
Simulation results under the null hypothesis. A) Empirical type I error rates with 95% CI at α = 0.05. B) Mean square error. **C)** Average causal estimation with 95% CI. Each column corresponds to scenarios with 30%, 50%, and 70% invalid IVs. Each row represents scenarios of balanced pleiotropy, directional pleiotropy, and InSIDE violation with θ = 0.1, 0.4, and 0.7, respectively, from top to bottom. Blue dashed lines indicate expected values. The sample size is 50,000. The number of SNPs is 100, all of which were initially provided to each method.

**Figure 3:**
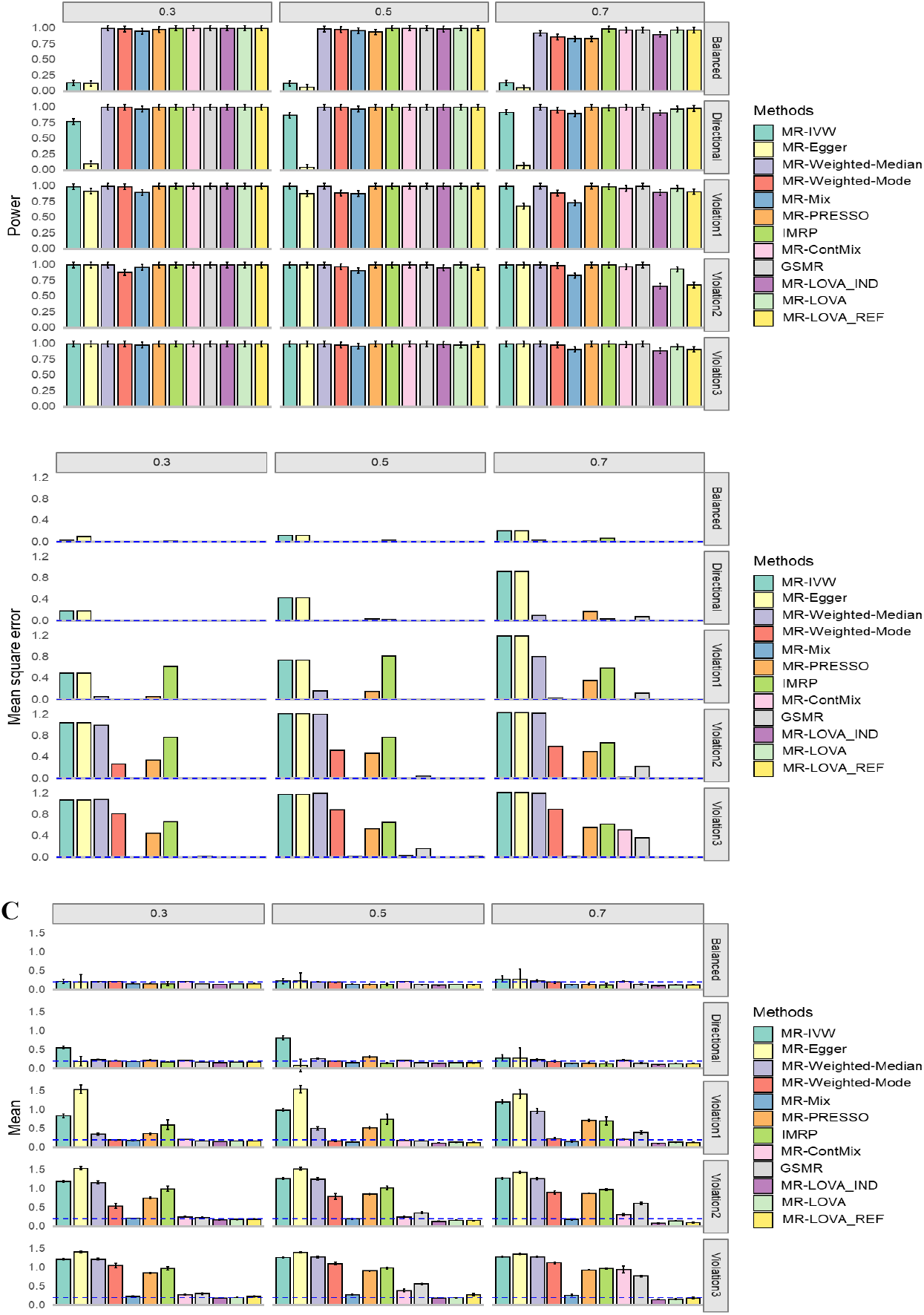
Simulation results under alternative hypothesis. A) Empirical Power with 95% CI. B) Mean square error. C) Average causal estimation with 95% CI. Each column corresponds to 30%, 50%, 70% invalid IVs. Each row corresponds to balanced pleiotropy, directional pleiotropy, and InSIDE violation with *θ* of 0.1, 0.4 and 0.7 respectively from top to bottom. The blue dash lines are expected values. The sample size is 50,000. The number of SNPs is 100, all of which were initially provided to each method.

In simulation scenario 3 (directional pleiotropy), increasing the proportion of invalid instruments similarly inflates overall type I error rates (Figure 2A) and increases MSE (Figure 2B), resembling scenario 2. Some MR methods exhibit significant bias (Figure 2C), such as IVW, MR-PRESSO, and weighted median. Nonetheless, our proposed method maintains minimal inflation of type I error rates and MSE and causal estimate remains unbiased even at 70% invalid instruments (Figure 2).

In simulation scenario 4 (directional pleiotropy + InSIDE violated), all methods demonstrate notable inflation in overall type I error rates (Figure 2A) and increased MSE (Figure 2B) as the proportion of invalid instruments increases. However, the proposed methods appeared to better control type I error rate (Figure 2A), with further improvement observed through permutation (Supplementary Figure 1). Most methods show substantial bias, particularly under conditions with large genetic effects associated with confounders and high proportions of invalid instruments (Figure 2B and C). In contrast, our proposed method exhibits only slight bias under these stringent conditions.

With a smaller sample size (Supplementary Figure 2) or a smaller number of causal variants (Supplementary Figure 3), the results remained consistent.

Under the alternative hypothesis (non-zero causal effects), the proposed method demonstrates adequate power with a sample size of 50,000, comparable to other established MR methods (Figure 3A). It is important to note that the inclusion of a permutation test in the proposed method may slightly reduce its power (Supplementary Figure 1). With a smaller sample size, our method shows lower power compared to other methods (Supplementary Figure 4), possibly due to their higher power driven by spurious inflation (Figure 3). However, across all scenarios (2–4), the proposed method consistently outperforms others in terms of MSE and bias (Figure 3B &C).

We also examined and compared the performance of the methods using all available SNPs (Figures 2 and 3) and SNPs selected with a p-value threshold of 5e-2 or 5e-08 from the exposure GWAS in the simulations (Supplementary Figures 5–8). This analysis aimed to assess the impact of strict versus relaxed relevance assumptions. We found that the performance of each method was mostly invariant, except for a decrease in power for the proposed method and a poorer MSE for IMRP with a stringent p-value threshold (i.e., fewer IVs selected). Notably, there was some improvement in MSE for MR-Weighted-Median, MR-Weighted-Mode, MR-PRESSO, GSMR, and MR-ConMix under specific conditions, such as the null hypothesis with 70% invalid instruments.

For the proposed methods, performance remains consistent regardless of whether individual-level data or summary-level data is used, or whether correlations between IVs are accounted for (see MR-LOVA_IND, MR-LOVA, and MR-LOVA_REF in Figures 1 – 3, Supplementary Figures 1 – 8). Therefore, we use MR-LOVA exclusively for downstream real data analysis.

Finally, we verified the proposed method’s ability to identify directional pleiotropy and violations of the InSIDE assumption using all available IVs (Supplementary Figures 11 and 12). Our method outperforms MR-Egger in its power to detect directional horizontal pleiotropy while both methods effectively control the type I error rate (Supplementary Figure 11). Additionally, our method proves effective in testing for violations of the InSIDE assumption (Supplementary Figure 12).

### Why does the MR-LOVA perform better?

As depicted in Figures 2 and 3, our proposed method consistently outperforms other methods, particularly evident when the InSIDE assumption is violated, with a proportion of invalid IVs at 70% and θ=0.7. Figure 4 illustrates that the estimated direct SNP effects on the outcome, i.e. **u** that are corrected for vertical pleiotropy, tend towards zero when using IVs selected by our proposed method. In contrast, other methods show a wider spread of effects, suggesting a violation of the exclusion restriction assumption–indicating IVs with significant direct effects on the outcome without a vertical pleiotropy pathway. The positively skewed distribution also indicates directional pleiotropy effects influencing the outcomes (Figure 4). Table 1 demonstrates that our method exhibits higher sensitivity and specificity in selecting valid IVs compared to MR-PRESSO, GSMR, MR-ConMix and IMRP, which are current methods available for disentangling valid and invalid variants.

**Table 1:** Evaluation of valid instrument selection by methods when the InSIDE assumption is violated, with 70% invalid IVs and θ=0.7.

|  | TP | FP | FN | TN | Sensitivity | Specificity | Accuracy | # selected IV |
| --- | --- | --- | --- | --- | --- | --- | --- | --- |
| MR-LOVA | 27.73 | 3.76 | 2.27 | 66.24 | 0.92 | 0.95 | 0.94 | 31.49 |
| IMRP | 11.02 | 17.59 | 18.98 | 52.41 | 0.37 | 0.75 | 0.63 | 28.61 |
| MR-ConMix | 7.45 | 11.74 | 13.92 | 36.53 | 0.25 | 0.25 | 0.44 | 19.19 |
| MR-PRESSO | 25.28 | 26.27 | 4.72 | 43.73 | 0.84 | 0.62 | 0.69 | 51.55 |
| GSMR | 18.35 | 20.49 | 11.65 | 49.51 | 0.61 | 0.71 | 0.68 | 38.84 |

**Figure 4:**
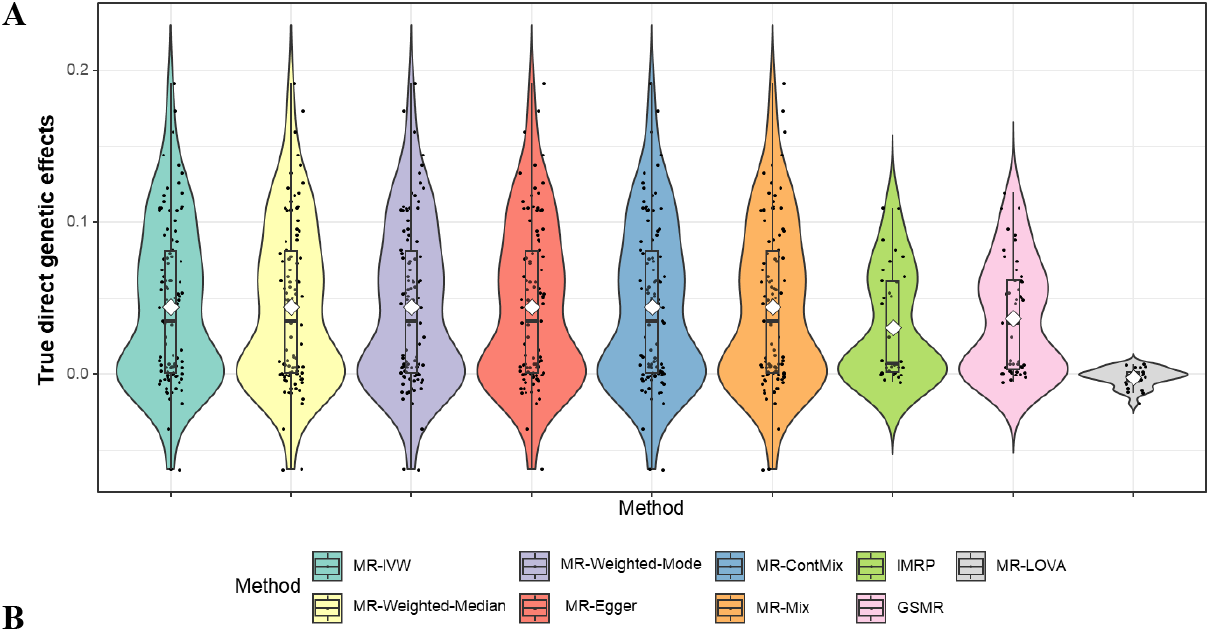
Distribution of direct genetic effects of IVs used in respective methods. The panel depicts a scenario where the InSIDE assumption is violated, with a proportion of invalid IVs at 70% and θ=0.7. The direct genetic effects (**u** in eq. (1)) were estimated for each method based on the IVs selected by that method, corrected for vertical pleiotropy using the true *τ*, and are presented in the plot.

This table evaluates the ability of MR-LOVA, IMRP, MR-PRESSO, GSMR, and MR-ConMix to select valid instruments. Results are based on the average of 100 iterations with a sample size of 50,000 and 100 instrumental variants.TP= true positive, the valid instrument correctly classified as valid by the MR methods. FP= false positive, the invalid instruments classified as valid. FN= false negative, valid instruments classified as invalid. TN = true negative, invalid instruments correctly identified by the methods.

### Real data analysis

The findings from our analysis examining the causal relationships between blood lipids, blood pressure, BMI, and their respective outcomes (CAD for blood lipids and blood pressure, and MetS for BMI and MetS score) are summarized in Table 2. We employed a range of MR methods, including MR-IVW, MR-Egger, MR-weighted-median, MR-weighted-mode, MR-PRESSO, MR-ConMix, IMRP, GSMR, and the proposed method, MR-LOVA.

**Table 2:** Estimates and 95% confidence intervals for causal effects and genetic correlation of various risk-factors on disease outcomes.

| Traits | parameters | MR-LOVA | IMRP | MR-ConMix | MR-IVW | MR-Egger | MR-weighted-mode | MR-weighted-median | MR-Mix | MR-PRESSO | GSMR | GC |
| --- | --- | --- | --- | --- | --- | --- | --- | --- | --- | --- | --- | --- |
| **LDL → CAD** | Estimate (SE) | 0.37(0.018) | 0.37(0.017) | 0.4(0.020) | 0.29(0.023) | 0.27(0.037) | 0.20(0.025) | 0.27(0.029) | 0.38(0.040) | 0.32(0.020) | 0.34(0.020) | 0.22(0.041) |
|  | P | 3.24E-41 | 3.33E-101 | 1.53E-27 | 6.33E-37 | 7.70E-14 | 8.07E-15 | 6.67E-21 | 1.08E-21 | 5.91E-35 | 1.48E-64 | 1.32E-07 |
|  | IVs (165) | 115 | 135 | 165 | 165 | 165 | 165 | 165 | 165 | 155 | 134 |  |
| **HDL → CAD** | Estimate (SE) | -0.03(0.021) | -0.02(0.021) | 0.00(0.041) | -0.15(0.035) | 0.05(0.068) | 0.03(0.054) | -0.05(0.038) | 0.02(0.121) | -0.14(0.028) | -0.19(0.027) | -0.24(0.036) |
|  | P | 1.49E-01 | 2.44E-01 | 1.00E+00 | 1.38E-05 | 5.06E-01 | 5.92E-01 | 1.66E-01 | 8.69E-01 | 8.18E-07 | 1.70E-12 | 1.69E-11 |
|  | IVs (153) | 103 | 104 | 153 | 153 | 153 | 153 | 153 | 153 | 139 | 105 |  |
| **SBP → CAD** | Estimate (SE) | 0.37(0.029) | 0.40(0.028) | 0.43(0.041) | 0.44(0.047) | 0.60(0.219) | 0.36(0.112) | 0.43(0.042) | 0.46(0.063) | 0.45(0.034) | 0.44(0.031) | 0.34 (0.029) |
|  | P | 1.42E-28 | 2.46E-46 | 1.33E-20 | 7.33E-21 | 6.04E-03 | 1.30E-03 | 1.50E-23 | 2.81E-13 | 1.48E-30 | 2.50E-46 | 8.60E-32 |
|  | IVs (288) | 218 | 228 | 288 | 288 | 288 | 288 | 288 | 288 | 268 | 205 |  |
| **DBP → CAD** | Estimate (SE) | 0.32(0.031) | 0.36(0.030) | 0.40(0.043) | 0.41(0.048) | 0.76(0.229) | 0.37(0.136) | 0.41(0.044) | 0.39(0.072) | 0.40(0.038) | 0.37(0.033) | 0.28 (0.0371) |
|  | P | 8.46E-20 | 2.54E-33 | 3.86E-14 | 5.84E-18 | 8.55E-04 | 6.60E-03 | 1.17E-20 | 5.94E-08 | 4.26E-22 | 5.64E-29 | 4.64E-14 |
|  | IVs (264) | 190 | 204 | 264 | 264 | 264 | 264 | 264 | 264 | 247 | 193 |  |
| **BMI → MetS** | Estimate (SE) | 0.46(0.018) | 0.47(0.016) | 0.49(0.015) | 0.43(0.029) | 0.45(0.081) | 0.44(0.033) | 0.44(0.029) | 0.48(0.027) | 0.45(0.020) | 0.47(0.02) | 0.65 (0.028) |
|  | P | 2.80E-31 | 3.73E-195 | 3.86E-36 | 4.92E-50 | 3.40E-08 | 1.06E-39 | 4.48E-52 | 4.45E-70 | 4.24E-36 | 8.83E-123 | 1.03E-121 |
|  | IVs (87) | 53 | 73 | 87 | 87 | 87 | 87 | 87 | 87 | 81 | 66 |  |
| **BMI → MetS score** | Estimate (SE) | 0.52(0.025) | 0.77(0.015) | 0.85(0.015) | 0.75(0.025) | 0.88(0.068) | 0.81(0.044) | 0.81(0.026) | 0.80(0.034) | 0.76(0.02) | 0.70 (0.019) | 0.81 (0.016) |
|  | P | 2.49E-15 | 0.00E+00 | 1.61E-104 | 8.58E-203 | 0.00E+00 | 6.16E-74 | 5.04E-213 | 1.17E-123 | 1.64E-54 | 2.44E-292 | 0 |
|  | IVs (90) | 22 | 64 | 90 | 90 | 90 | 90 | 90 | 90 | 86 | 66 |  |

CAD: coronary artery disease; MetS: metabolic syndrome; MetS score: metabolic syndrome score; LDL: low-density lipoprotein cholesterol; HDL: high-density lipoprotein cholesterol; DBP: diastolic blood pressure; SBP: systolic blood pressure, BMI: body mass index. GC: genetic correlation estimated using LD score regression. IVs: numbers of independent SNPs which reach genome-wide significance in the study associated with exposure (clumped based on 500 kb window sizes and $r^2$ threshold of 0.1)

For LDL vs. CAD, all methods identified significant associations, with MR-LOVA, IMRP, MR-ConMix, MR-Mix, and GSMR providing the most consistent estimates, ranging from 0.34 to 0.40. For HDL vs. CAD, most methods did not find a significant association, except for MR-IVW, MR-PRESSO, and GSMR, which detected weak but significant effects. For blood pressure vs, CAD, consistent positive associations were observed across all methods for both systolic and diastolic blood pressure, confirming a robust relationship between blood pressure and CAD.

In contrast to CAD for blood lipids and blood pressure (genetic correlations ranging from -0.23 to 0.34), MetS for BMI and MetS score showed higher genetic correlations (0.65 to 0.81) (Table 2). For BMI vs. MetS, the analysis consistently supported a causal effect of BMI on MetS across most methods. Specifically, MR-LOVA showed a strong and significant association with MetS (*τ* = 0.46, SE = 0.018), suggesting that higher BMI causally increases the risk of MetS. Other methods yielded similar estimates, ranging from 0.43 (SE = 0.029) for MR-IVW to 0.49 (SE = 0.015) for MR-ConMix.

However, when using MetS score as the outcome, the proposed method estimated 0.52 (SE 0.025), whereas other existing methods provided substantially higher estimates, ranging from 0.70 (SE 0.025) for GSMR to 0.88 (SE 0.068) for MR-Egger. It is noted that the estimated genetic correlation between MetS score and BMI are notably higher (Table 1), indicating persistent horizontal pleiotropy.

In line with our simulation study (Figure 4), we compared the inferred direct genetic effects across methods. Figure 5 illustrates that the estimated direct SNP effects on the outcome from the proposed method tend towards zero in the absence of vertical pleiotropy effects, contrasting other methods that show a wider spread of effects, indicating the violation of exclusion restriction assumption. This result was invariant when using various estimated causal effects from other methods in the inference of the direct SNP effects on the outcome (Supplementary Figure 9 & 10).

**Figure 5:**
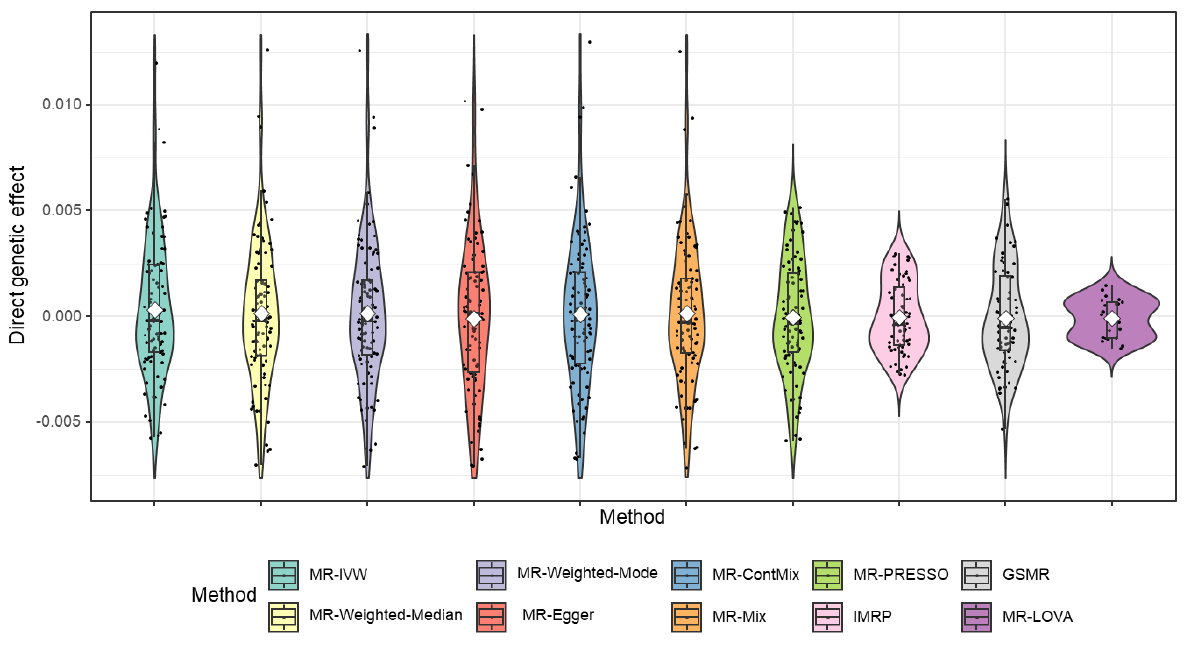
The distribution of the direct genetic effect of BMI’s IVs on MetS score estimated based on the causal effect estimated by respective methods. The direct effects were inferred from Eq. 2 using the estimated causal effects for each method. The direct genetic effects (u in eq. (1)) were estimated for each method based on the IVs selected by that method, corrected for vertical pleiotropy using the *τ* estimated by the method, and are presented in the plot.

There was no evidence of a positively skewed distribution, indicating the absence of directional pleiotropy effects. Similarly, the novel statistical test for directional pleiotropy showed non-significant for all analyses except LDL vs. CAD. The InSIDE assumption tests were also not significant in all cases except for BMI and MetS scores (Supplementary Table 1).

## Discussion

Our study explores the application of MR methods to understand the genetic underpinnings of complex traits influenced by both genetic and environmental factors. The interplay of these factors, often characterized by horizontal and vertical pleiotropy, presents challenges in accurately estimating causal effects^(20, 41)^. Our proposed method, MR-LOVA, addresses these challenges by introducing a novel approach to MR analysis. Unlike conventional methods that assume a mixture model for residuals based on IV validity, MR-LOVA utilizes latent phenotypes of the outcome variable that explicitly exclude vertical pleiotropy effects. This method iteratively refines estimates through an EM algorithm, enhancing precision in causal effect estimation.

In our simulation studies, we systematically evaluated MR-LOVA alongside established MR methods across various scenarios. Under scenarios of balanced pleiotropy, directional pleiotropy, and violations in InSIDE assumption^(19)^, MR-LOVA consistently demonstrated minimal inflation of type I error rates and MSE, indicating robust performance in controlling false positives and maintaining accuracy in causal effect estimates. These findings underscore the method’s ability to handle complex genetic architectures and mitigate biases that may arise from pleiotropic effects. Comparatively, widely used MR methods such as IVW, MR-Egger, MR-weighted-median and MR-weighted-mode ^(19, 20, 24)^ exhibited varying degrees of bias and inflated error rates under scenarios with invalid instruments or directional pleiotropy. This highlights the importance of methodological advancements like MR-LOVA, which offer improved bias control and robustness in the presence of genetic confounding.

MR-PRESSO, GSMR, and IMRP^(15, 16, 18)^ use heterogeneity tests to detect pleiotropic variants among the IVs and provide global tests to assess the overall presence of pleiotropic effects. While these methods are useful for identifying and removing outliers, they do not differentiate between types of pleiotropy, such as balanced, directional, or violations of the InSIDE assumption. In contrast, the InSIDE assumption violation test implemented in MR-LOVA specifically addresses violations of the InSIDE assumption, distinguishing them from balanced and directional pleiotropy. This distinction is crucial as violations of the InSIDE assumption generally pose a greater concern for method performance. Additionally, while MR-Egger^(12)^ indirectly tests for directional pleiotropy through the intercept, MR-LOVA offers a more direct assessment, providing improved statistical power compared to MR-Egger.

When a genetic variant *i* is not a valid IV and has a direct genetic effect on the outcome (u_*i*_≠0), the causal effect for variant *i* can be expressed as *τ*_*i*_=*τ*+*u*_*i*_/*b*_*i*_, where the latter term represents the bias factor^(12)^. The smaller the value of *bi* (the genetic effect on the exposure), the larger the bias term, indicating weak IV bias. However, as illustrated in Figures 4 and 5, MR-LOVA explicitly assesses the direct effect on the outcome and excludes IVs with a latent outcome variable GWAS p-value < 0.05. This suggests that weak IV bias is less of a concern in this MR-LOVA analysis compared to other MR methods. Additionally, the p-value threshold for the latent outcome variable GWAS in MR-LOVA can be made more stringent (e.g., < 0.5) to further minimize weak IV bias, highlighting a unique property of MR-LOVA.

In conclusion, MR-LOVA represents a significant advancement in MR methodology, offering researchers a more reliable tool for causal inference in studies of complex traits. Notably, using MR-LOVA, we uncovered the causal effects of BMI on MetS and on a composite MetS score—findings that might have been distorted using conventional MR methods due to the violation of the InSIDE assumption and the high genetic correlation between BMI and MetS score. Future research could further explore its application across diverse populations and in different phenotypic contexts, potentially uncovering novel insights into the genetic basis of complex diseases and traits.

## Data Availability

All data produced in the present study are available upon reasonable request to the authors.

https://github.com/lamessad/MRLOVA

## Code availability

The proposed methods are implemented in R package MRLOVE, which is publicly available to download on GitHub at https://github.com/lamessad/MRLOVA. All other MR methods used for comparison are in publicly available R packages with links given in the web resources section.

## AUTHOR CONTRIBUTIONS

S. Hong Lee conceived the idea, derived the proposed Mendelian randomization (MR) models, and supervised the study. Lamessa Amente contributed to performing simulations, data extraction, data analysis, and visualizations. Both Lamessa Amente and S. Hong Lee wrote the first draft of the manuscript. S. Hong Lee initially created the computer code for simulations and analyses and reviewed the simulations, data extraction, data analysis, and visualizations throughout the study. Lamessa Amente and S. Hong Lee also worked on preparing the R package. Thuc Lee, Natalie Mills, and Elina Hyppönen reviewed the manuscript and provided critical feedback and suggestions. All authors discussed the results and contributed to finalizing the manuscript.

## ACKNOWLEDGMENTS

Lamessa Amente acknowledges funding from the Australian Government Research Training Program (RTP) scholarship. The authors would like to thank staff and participants of the Atherosclerosis Risk in Communities Study (phs000280) and the UK Biobank for their important contributions to the GWAS summary statistics. The Atherosclerosis Risk in Communities Study has been funded in whole or in part with federal funds from the National Heart, Lung, and Blood Institute, National Institutes of Health, Department of Health and Human Services (under contract numbers HHSN268201700001I, HHSN268201700002I, HHSN268201700003I, HHSN268201700005I, HHSN268201700004I). The analyses were performed using computational resources provided by the Australian Government through Gadi under the National Computational Merit Allocation Scheme (NCMAS) and under University of South Australia (UniSA), and HPCs (Statgen and Statgen 2 servers) managed by UniSA IT. We thank University of South Australia IT team for their support in accessing the servers. Finally, we would like to thank the Statistical Genetics Group at the Australian Centre for Precision Health for helpful discussions contributing to this work.

## Web resources

https://github.com/MRCIEU/TwoSampleMR

https://cran.r-project.org/web/packages/MendelianRandomization

https://github.com/rondolab/MR-PRESSO

https://github.com/XiaofengZhuCase/IMRP

https://github.com/gqi/MRMix

## References

1. Dempfle A, Scherag A, Hein R, Beckmann L, Chang-Claude J, Schafer H. Gene-environment interactions for complex traits: definitions, methodological requirements and challenges. Eur J Hum Genet. 2008;16(10):1164–72.

2. Burgess S, Butterworth A, Thompson SG. Mendelian randomization analysis with multiple genetic variants using summarized data. Genet Epidemiol. 2013;37(7):658–65.

3. Davies NM, Holmes MV, Davey Smith G. Reading Mendelian randomisation studies: a guide, glossary, and checklist for clinicians. BMJ. 2018;362:k601.

4. Sivakumaran S, Agakov F, Theodoratou E, Prendergast JG, Zgaga L, Manolio T, et al. Abundant pleiotropy in human complex diseases and traits. Am J Hum Genet. 2011;89(5):607–18.

5. Burgess S, Davey Smith G, Davies NM, Dudbridge F, Gill D, Glymour MM, et al. Guidelines for performing Mendelian randomization investigations: update for summer 2023. Wellcome Open Res. 2019;4:186.

6. Solovieff N, Cotsapas C, Lee PH, Purcell SM, Smoller JW. Pleiotropy in complex traits: challenges and strategies. Nat Rev Genet. 2013;14(7):483–95.

7. Lee SH, Yang J, Goddard ME, Visscher PM, Wray NR. Estimation of pleiotropy between complex diseases using single-nucleotide polymorphism-derived genomic relationships and restricted maximum likelihood. Bioinformatics. 2012;28(19):2540–2.

8. Pierce BL, Ahsan H, Vanderweele TJ. Power and instrument strength requirements for Mendelian randomization studies using multiple genetic variants. Int J Epidemiol. 2011;40(3):740–52.

9. Yao M, Miller GW, Vardarajan BN, Baccarelli AA, Guo Z, Liu Z. Selecting Valid Genetic Instruments and Constructing Robust Confidence Intervals for Two-sample Mendelian Randomization Using Genome-wide Summary Statistics. 2023.

10. Sanderson E, Davey Smith G, Windmeijer F, Bowden J. An examination of multivariable Mendelian randomization in the single-sample and two-sample summary data settings. Int J Epidemiol. 2019;48(3):713–27.

11. Burgess S, Foley CN, Allara E, Staley JR, Howson JMM. A robust and efficient method for Mendelian randomization with hundreds of genetic variants. Nat Commun. 2020;11(1):376.

12. Bowden J, Davey Smith G, Burgess S. Mendelian randomization with invalid instruments: effect estimation and bias detection through Egger regression. Int J Epidemiol. 2015;44(2):512–25.

13. Hartwig FP, Davey Smith G, Bowden J. Robust inference in summary data Mendelian randomization via the zero modal pleiotropy assumption. Int J Epidemiol. 2017;46(6):1985–98.

14. Bowden J, Davey Smith G, Haycock PC, Burgess S. Consistent Estimation in Mendelian Randomization with Some Invalid Instruments Using a Weighted Median Estimator. Genet Epidemiol. 2016;40(4):304–14.

15. Verbanck M, Chen CY, Neale B, Do R. Detection of widespread horizontal pleiotropy in causal relationships inferred from Mendelian randomization between complex traits and diseases. Nat Genet. 2018;50(5):693–8.

16. Zhu Z, Zheng Z, Zhang F, Wu Y, Trzaskowski M, Maier R, et al. Causal associations between risk factors and common diseases inferred from GWAS summary data. Nat Commun. 2018;9(1):224.

17. Qi G, Chatterjee N. Mendelian randomization analysis using mixture models for robust and efficient estimation of causal effects. Nat Commun. 2019;10(1):1941.

18. Zhu X, Li X, Xu R, Wang T. An iterative approach to detect pleiotropy and perform Mendelian Randomization analysis using GWAS summary statistics. Bioinformatics. 2021;37(10):1390–400.

19. Slob EAW, Burgess S. A comparison of robust Mendelian randomization methods using summary data. Genet Epidemiol. 2020;44(4):313–29.

20. Xue H, Shen X, Pan W. Constrained maximum likelihood-based Mendelian randomization robust to both correlated and uncorrelated pleiotropic effects. Am J Hum Genet. 2021;108(7):1251–69.

21. Zhu X. Mendelian randomization and pleiotropy analysis. Quant Biol. 2021;9(2):122–32.

22. Stephens M. A unified framework for association analysis with multiple related phenotypes. PLoS One. 2013;8(7):e65245.

23. Momin MM, Lee S, Wray NR, Lee SH. Significance tests for R2 of out-of-sample prediction using polygenic scores. Am J Hum Genet. 2023;110(2):349–58.

24. Lin Z, Deng Y, Pan W. Combining the strengths of inverse-variance weighting and Egger regression in Mendelian randomization using a mixture of regressions model. PLoS Genet. 2021;17(11):e1009922.

25. Nikpay M, Goel A, Won HH, Hall LM, Willenborg C, Kanoni S, et al. A comprehensive 1,000 Genomes-based genome-wide association meta-analysis of coronary artery disease. Nat Genet. 2015;47(10):1121–30.

26. Willer CJ, Schmidt EM, Sengupta S, Peloso GM, Gustafsson S, Kanoni S, et al. Discovery and refinement of loci associated with lipid levels. Nat Genet. 2013;45(11):1274–83.

27. Neale B. Rapid GWAS of thousands of phenotypes for 337,000 samples in the UK Biobank. https://www.nealelab.is/blog/2017/7/19/rapid-gwas-of-thousands-of-phenotypes-for-337000-samples-in-the-uk-biobank

28. Mounier N, Kutalik Z. Bias correction for inverse variance weighting Mendelian randomization. Genet Epidemiol. 2023;47(4):314–31.

29. Yengo L, Sidorenko J, Kemper KE, Zheng Z, Wood AR, Weedon MN, et al. Meta-analysis of genome-wide association studies for height and body mass index in approximately 700000 individuals of European ancestry. Hum Mol Genet. 2018;27(20):3641–9.

30. Lind L. Genome-Wide Association Study of the Metabolic Syndrome in UK Biobank. Metab Syndr Relat Disord. 2019;17(10):505–11.

31. van Walree ES, Jansen IE, Bell NY, Savage JE, de Leeuw C, Nieuwdorp M, et al. Disentangling Genetic Risks for Metabolic Syndrome. Diabetes. 2022;71(11):2447–57.

32. Amente LD, Mills NT, L. TD, Hypponen E, Lee SH. Unraveling phenotypic variance in metabolic syndrome through multi-omics. Hum Genet. 2024;143(1):35–47.

33. Bluher M. The distinction of metabolically ‘healthy’ from ‘unhealthy’ obese individuals. Curr Opin Lipidol. 2010;21(1):38–43.

34. Despres JP. Is visceral obesity the cause of the metabolic syndrome? Ann Med. 2006;38(1):52–63.

35. National Heart L, and Blood Institute. Metabolic syndrome: Causes. n.d.

36. Grundy SM. Obesity, metabolic syndrome, and cardiovascular disease. J Clin Endocrinol Metab. 2004;89(6):2595–600.

37. Alberti KG, Eckel RH, Grundy SM, Zimmet PZ, Cleeman JI, Donato KA, et al. Harmonizing the metabolic syndrome: a joint interim statement of the International Diabetes Federation Task Force on Epidemiology and Prevention; National Heart, Lung, and Blood Institute; American Heart Association; World Heart Federation; International Atherosclerosis Society; and International Association for the Study of Obesity. Circulation. 2009;120(16):1640–5.

38. Grotzinger AD, Rhemtulla M, de Vlaming R, Ritchie SJ, Mallard TT, Hill WD, et al. Genomic structural equation modelling provides insights into the multivariate genetic architecture of complex traits. Nat Hum Behav. 2019;3(5):513–25.

39. Graziano F, Grassi M, Bonati MT, Zanchetti A, Biino G. External validation of the MetS score, a prediction tool for metabolic syndrome. Nutr Metab Cardiovasc Dis. 2016;26(4):359–60.

40. Bulik-Sullivan B, Finucane HK, Anttila V, Gusev A, Day FR, Loh PR, et al. An atlas of genetic correlations across human diseases and traits. Nat Genet. 2015;47(11):1236–41.

41. Morrison J, Knoblauch N, Marcus JH, Stephens M, He X. Mendelian randomization accounting for correlated and uncorrelated pleiotropic effects using genome-wide summary statistics. Nat Genet. 2020;52(7):740–7.

