## Supplementary file for "A latent outcome variable approach for Mendelian randomization using the expectation maximization algorithm"

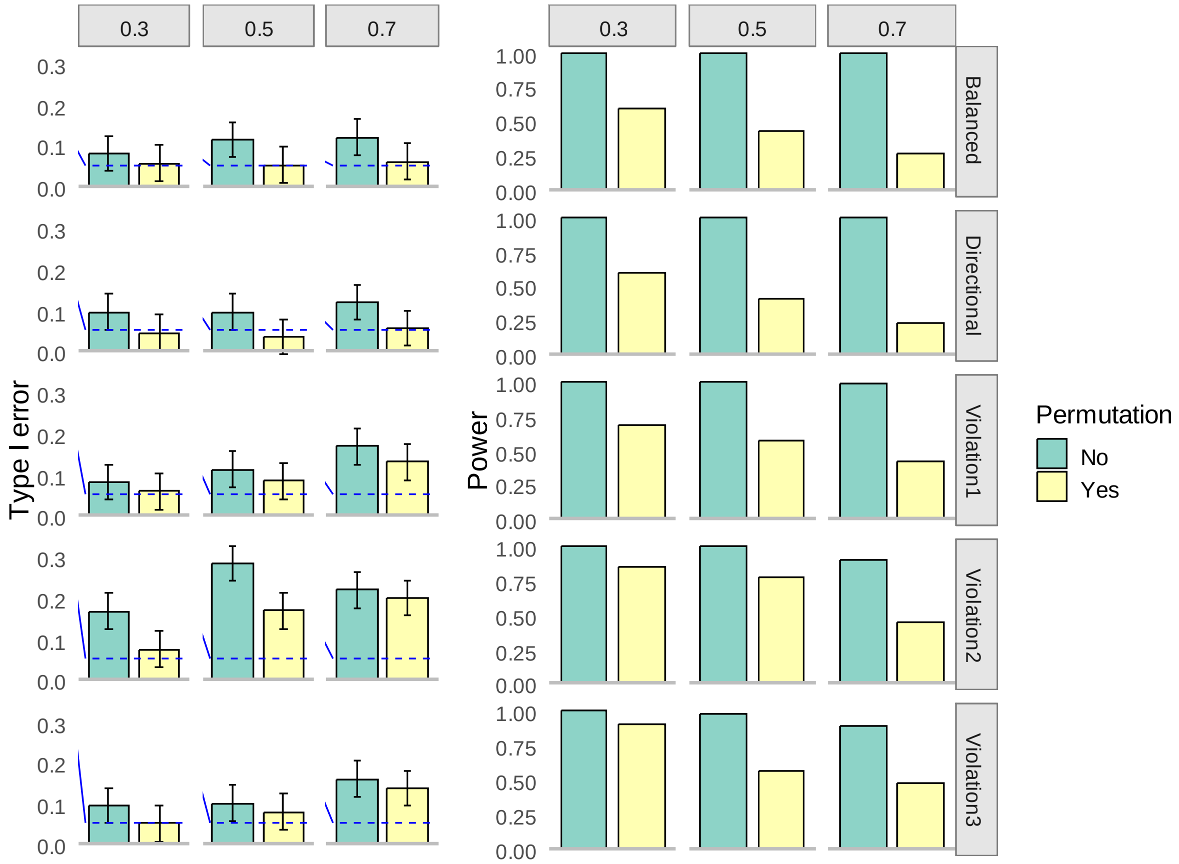


**Supplementary Figure 1: Permutation better controls type I error rates under null hypothesis (the left panel) and reduce power under alternative hypothesis (the right panel).** Each column corresponds to scenarios with 30%, 50%, and 70% invalid instrumental variables (IVs). Each row represents scenarios of balanced pleiotropy, directional pleiotropy, and InSIDE violation with $\theta$ = 0.1, 0.4, and 0.7, respectively, from top to bottom. Blue dashed lines indicate expected values. The sample size is 50,000. The number of SNPs is 100, all of which were initially provided to each method. The number of permutations applied is 1000.

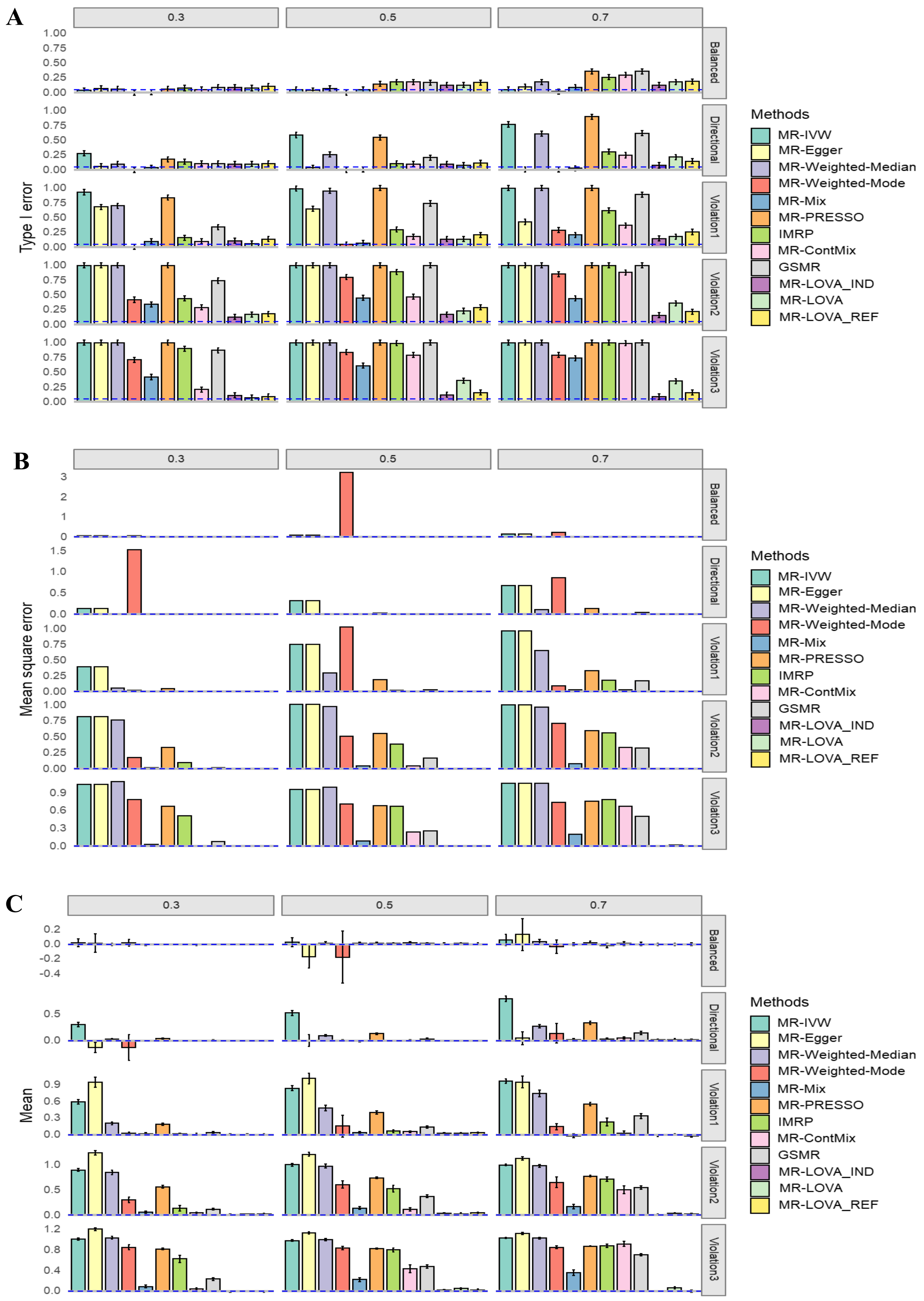


**Supplementary Figure 2: Simulation results under the null hypothesis. A)** Empirical type I error rates with 95% CI at α = 0.05**. B**) Mean square error**. C**) Average causal estimation with 95% CI. Each column corresponds to scenarios with 30%, 50%, and 70% invalid instrumental variables (IVs). Each row represents scenarios of balanced pleiotropy, directional pleiotropy, and InSIDE violation with $\theta$ = 0.1, 0.4, and 0.7, respectively, from top to bottom. Blue dashed lines indicate expected values. The sample size is 20,000. The number of SNPs is 100, all of which were initially provided to each method.

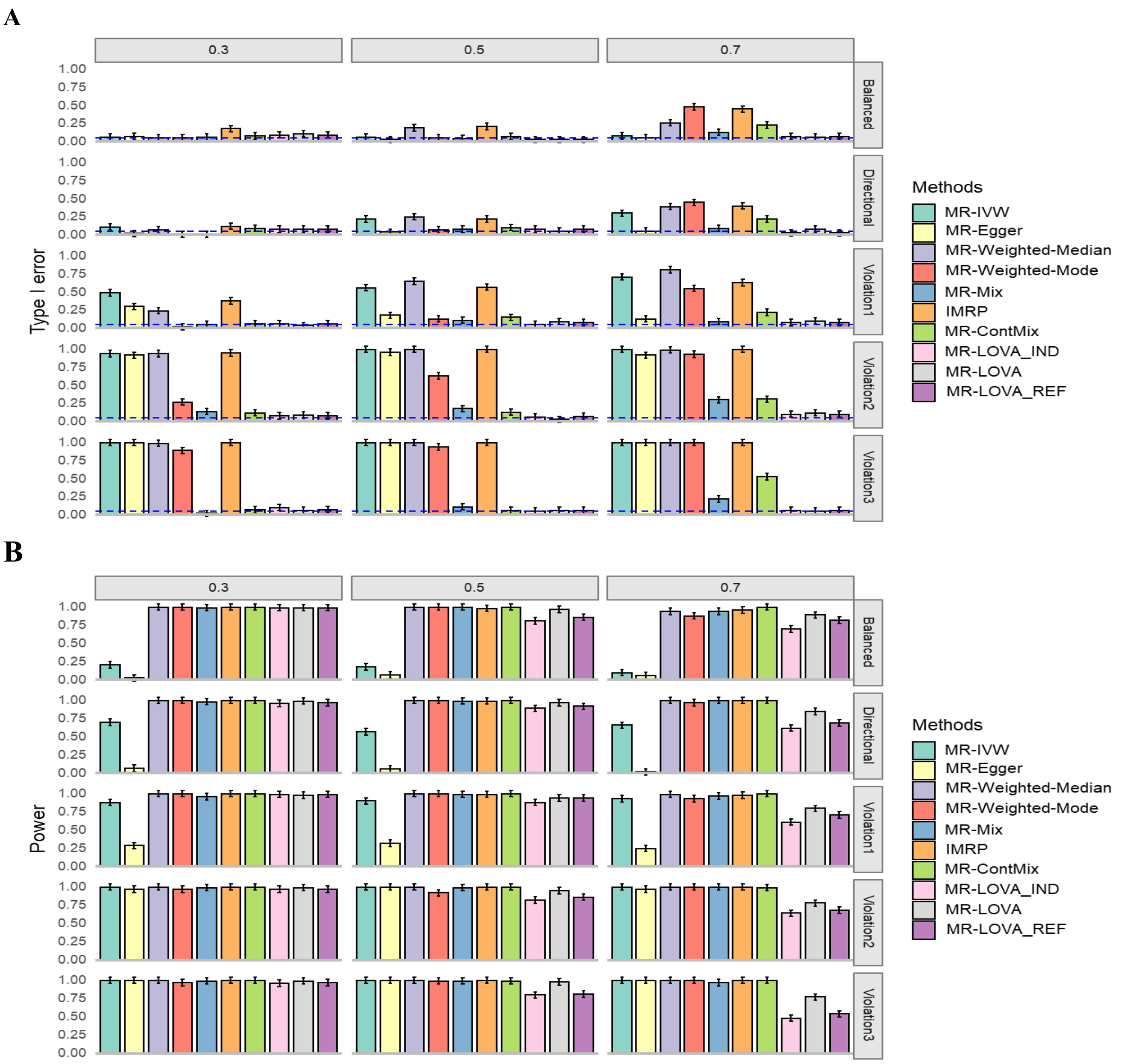


**Supplementary Figure 3:** Empirical type I error and power. **A**) Empirical type I error rates with 95% CI at $\alpha = 0.05$. **B**) Power under alternative hypothesis. Each column corresponds to scenarios with 30%, 50%, and 70% invalid IVs. Each row represents scenarios of balanced pleiotropy, directional pleiotropy, and InSIDE violation with $\theta$ = 0.1, 0.4, and 0.7, respectively, from top to bottom. Blue dashed lines indicate expected values. The sample size is 50,000. The number of SNPs is 30, all of which were initially provided to each method. MR-PRESSO and GSMR were excluded because of NA reported during simulation for same iterations.

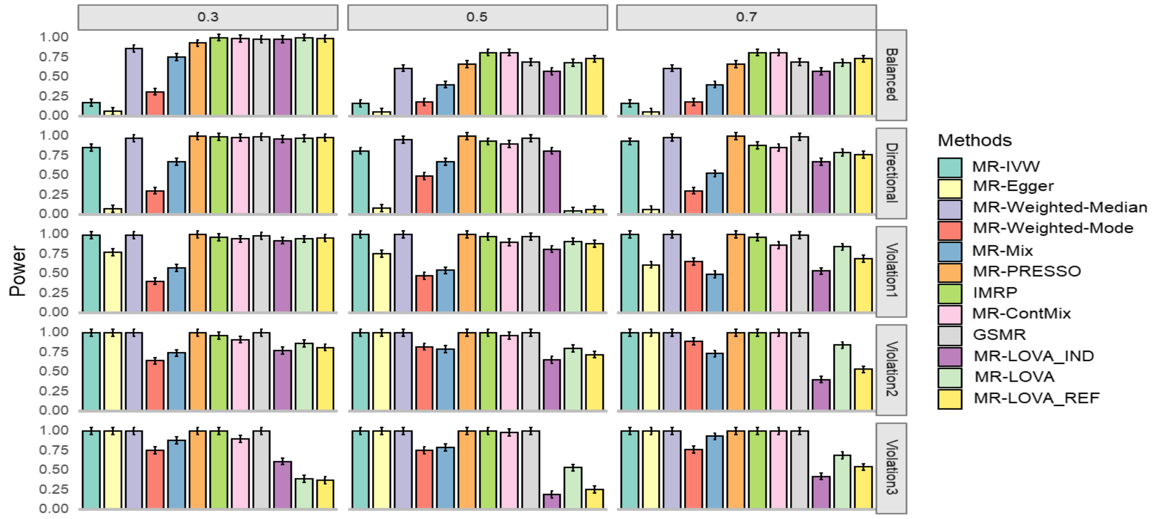


**Supplementary Figure 4: Simulation results under alternative hypothesis.** Empirical power with 95% CI. Each column corresponds to 30%, 50%, 70% invalid IVs. Each row corresponds to balanced pleiotropy, directional pleiotropy, and InSIDE violation with ɵ of 0.1, 0.4 and 0.7 respectively from top to bottom. The sample size is 20,000. The number of SNPs is 100, all of which were initially provided to each method.

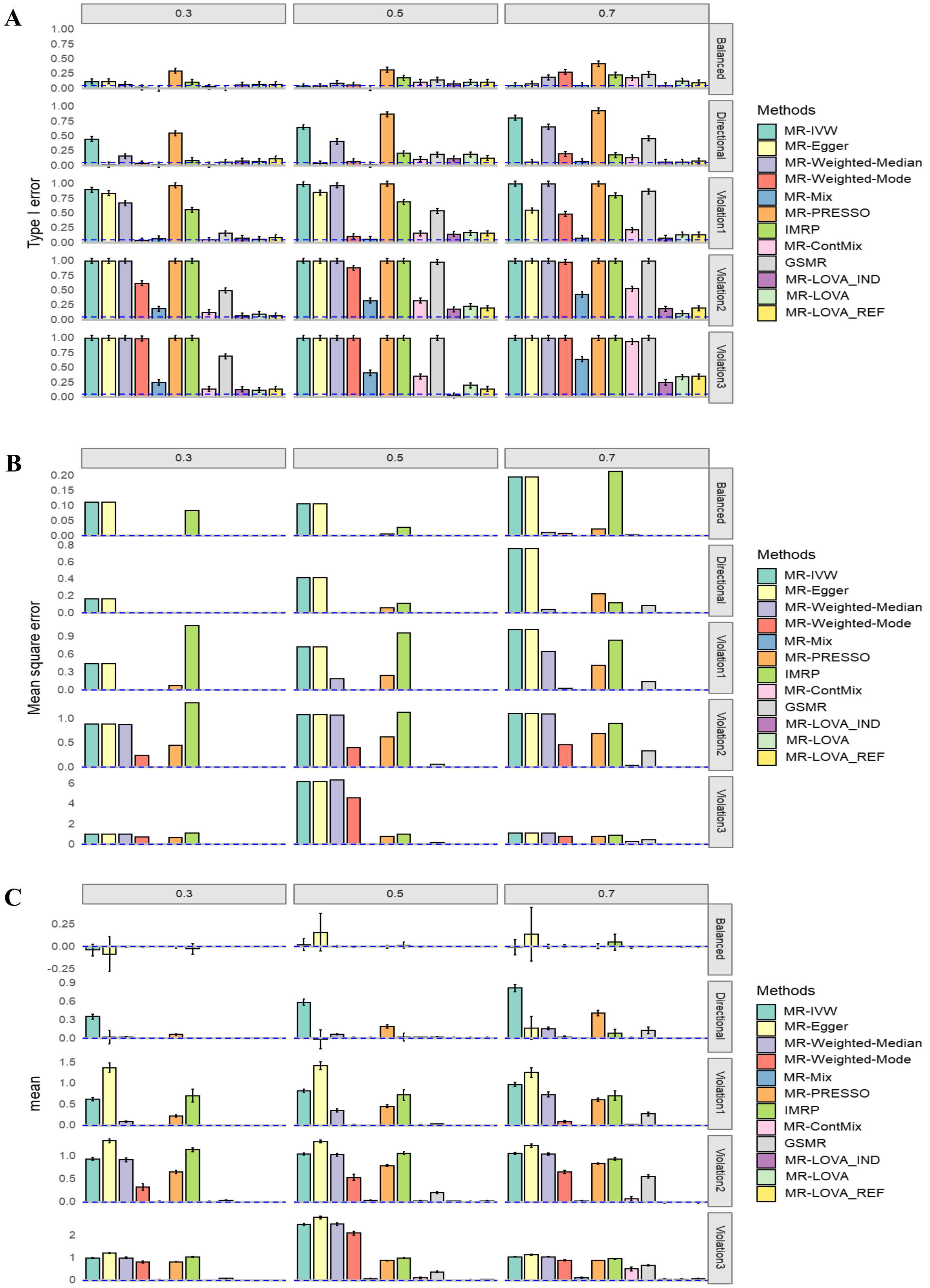


**Supplementary Figure 5: Simulation results under the null hypothesis.** A) Empirical Type I Error Rates with 95% CI at $\alpha = 0.05$. B) Mean Square Error. C) Average Causal Estimation with 95% CI. Each column corresponds to scenarios with 30%, 50%, and 70% invalid instrumental variables (IVs). Each row represents scenarios of balanced pleiotropy, directional pleiotropy, and InSIDE violation with $\theta$ = 0.1, 0.4, and 0.7, respectively, from top to bottom. Blue dashed lines indicate expected values. The sample size is 50,000. A p-value threshold < 5e-2 was applied to the exposure GWAS to select IVs.

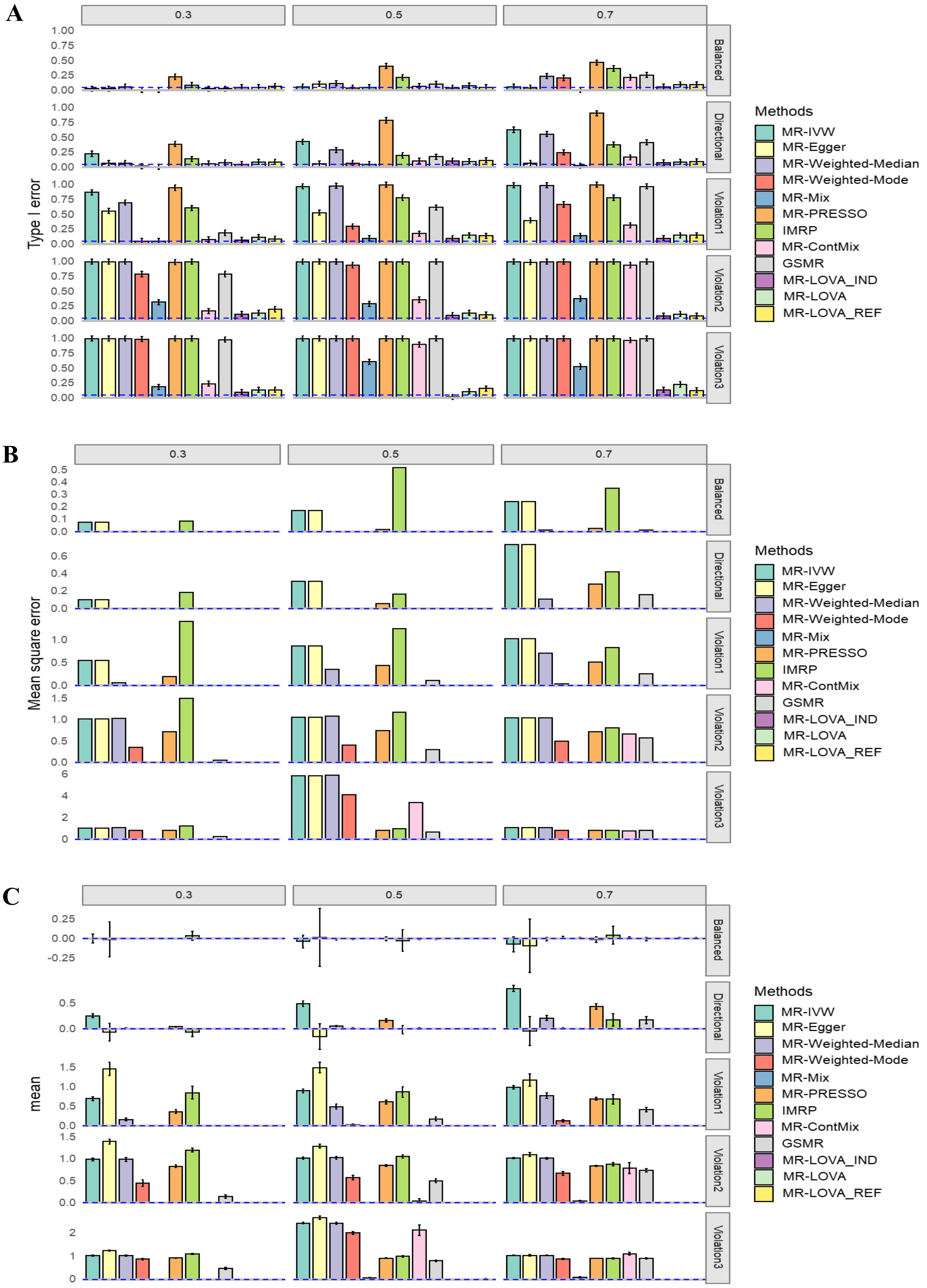


**Supplementary Figure 6: Simulation results under the null hypothesis.** A: Empirical type I error rates with 95% CI at α = 0.05. B: Mean square error. C: Average causal estimation with 95% CI. Each column corresponds to scenarios with 30%, 50%, and 70% invalid instrumental variables (IVs). Each row represents scenarios of balanced pleiotropy, directional pleiotropy, and InSIDE violation with θ = 0.1, 0.4, and 0.7, respectively, from top to bottom. Blue dashed lines indicate expected values. The sample size is 50,000. A p-value threshold < 5e-8 was applied to the exposure GWAS to select IVs.

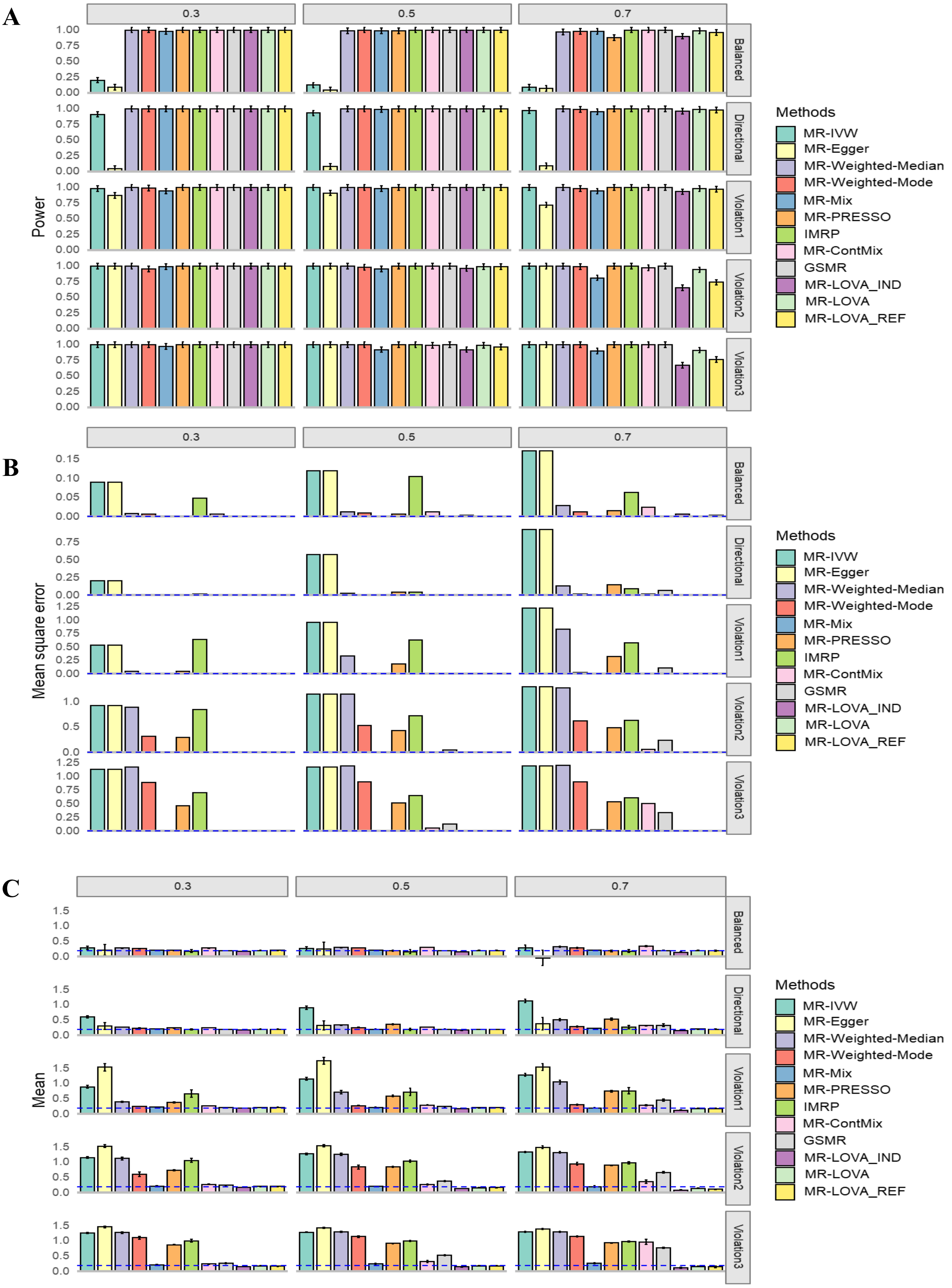


**Supplementary Figure 7: Simulation results under alternative hypothesis.** A) Empirical Power with 95% CI. B) Mean Square Error. C) Average Causal Estimation with 95% CI. Each column corresponds to 30%, 50%, 70% invalid IVs. Each row corresponds to balanced pleiotropy, directional pleiotropy, and InSIDE violation with ɵ of 0.1, 0.4 and 0.7 respectively from top to bottom. The blue dash lines are expected values. The sample size is 50,000. A p-value threshold < 5e-2 was applied to the exposure GWAS to select IVs.

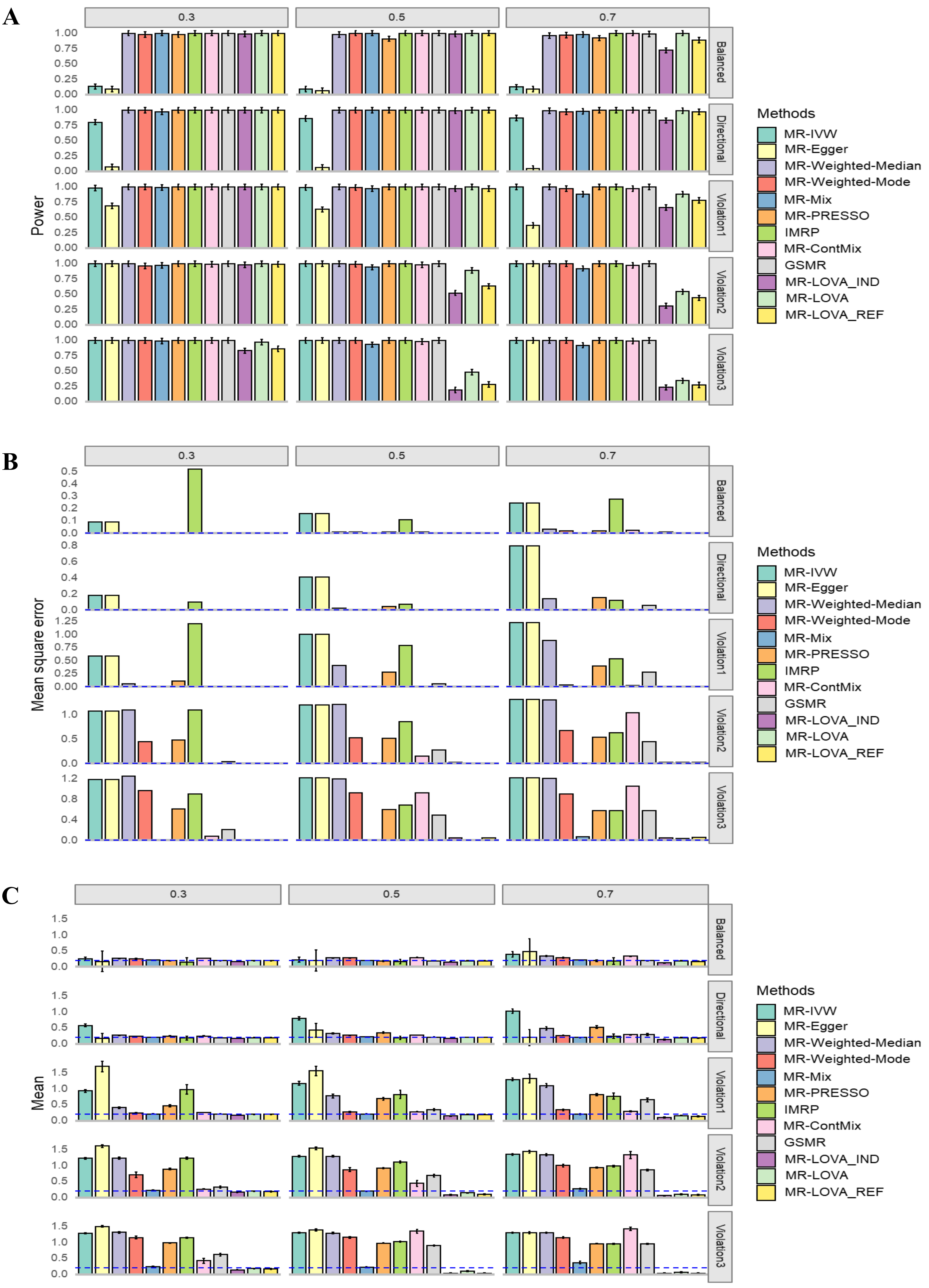


**Supplementary Figure 8: Simulation results under alternative hypothesis.** A) Empirical Power with 95% CI. B) Mean Square Error. C) Average Causal Estimation with 95% CI. Each column corresponds to 30%, 50%, 70% invalid IVs. Each row corresponds to balanced pleiotropy, directional pleiotropy, and InSIDE violation with ɵ of 0.1, 0.4 and 0.7 respectively from top to bottom. The blue dash lines are expected values. The sample size is 50,000. A p-value threshold < 5e-8 was applied to the exposure GWAS to select IVs.

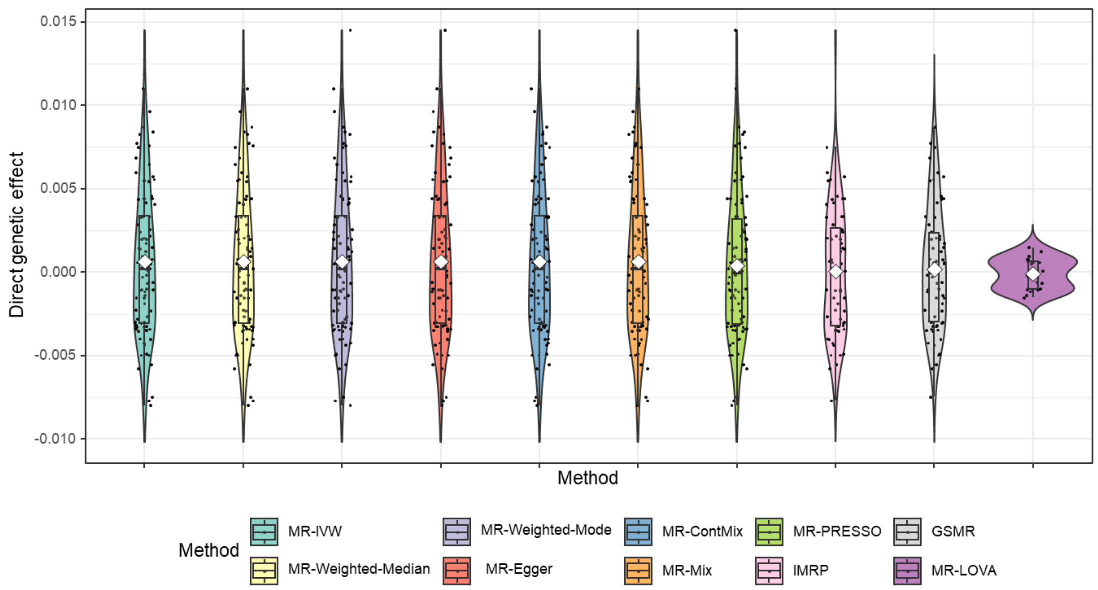


**Supplementary Figure 9: The distribution of the direct genetic effect of BMI's IVs on MetS score estimated based on the causal effect estimated by MR-LOVA.** The direct SNP effects (**u**) were inferred from Eq.1 using the estimated causal effects for each method.

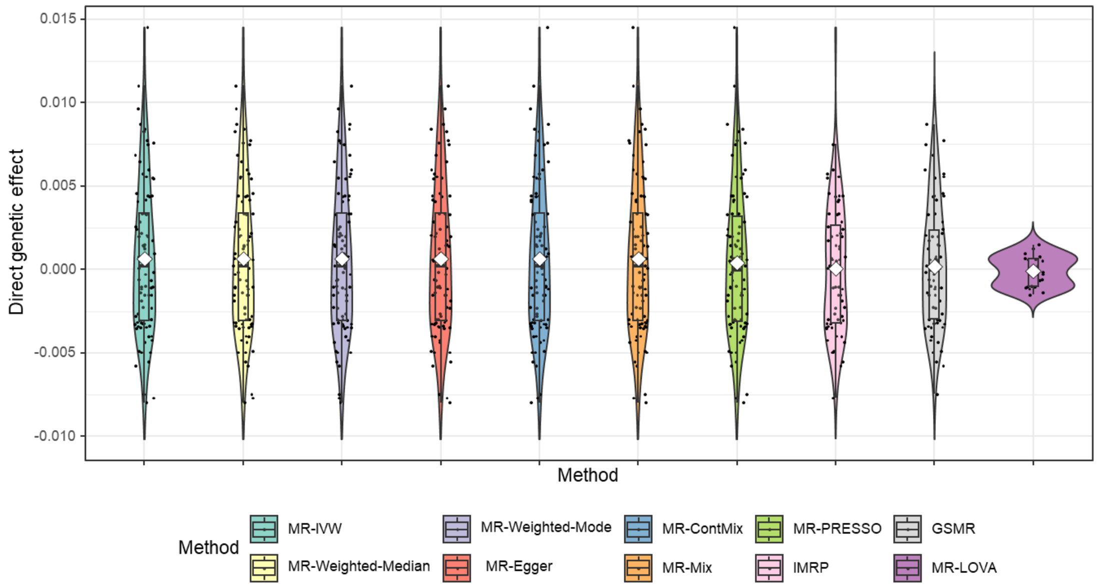


**Supplementary Figure 10: The distribution of the direct genetic effect of BMI's IVs on MetS score estimated based on the causal effect estimated by MR-Egger.** The direct effects were inferred from Eq. 1 using the estimated causal effects for each method.

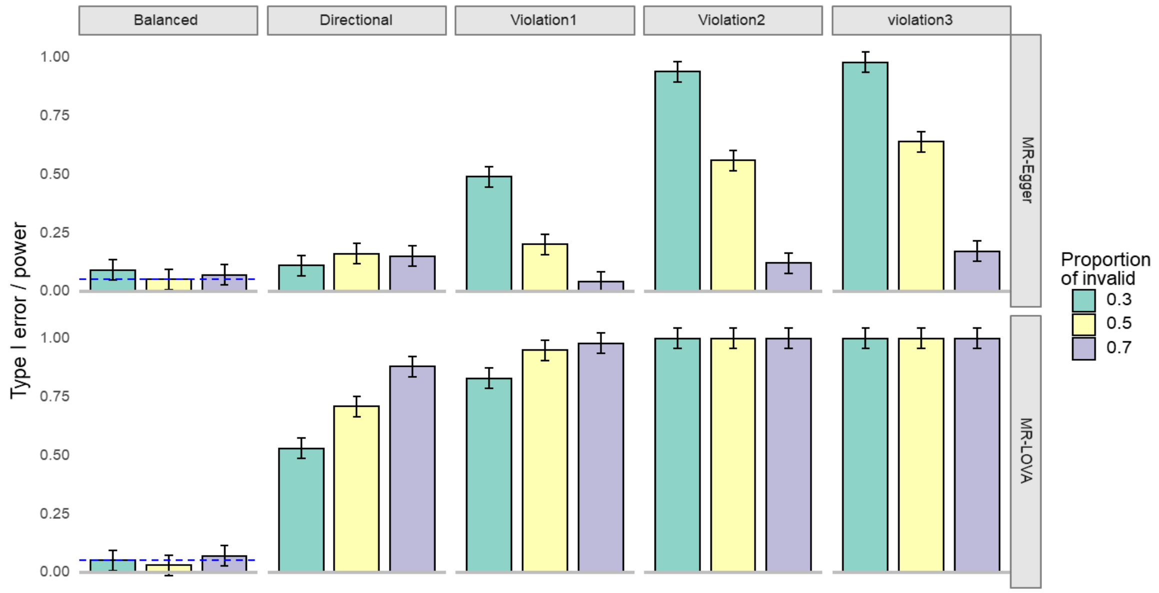


**Supplementary Figure 11: comparison of Type I error and Power of the directional pleiotropy test of MR-LOVA and MR-Egger.** The first column (i.e., balance) is simulated under no directional pleiotropy (i.e., type I error), while the last four columns are simulated under directional pleiotropy ('i.e., power). The blue dash lines are expected values for type I error. The sample size is 50,000. The number of SNPs is 100, all of which were initially provided to both methods.

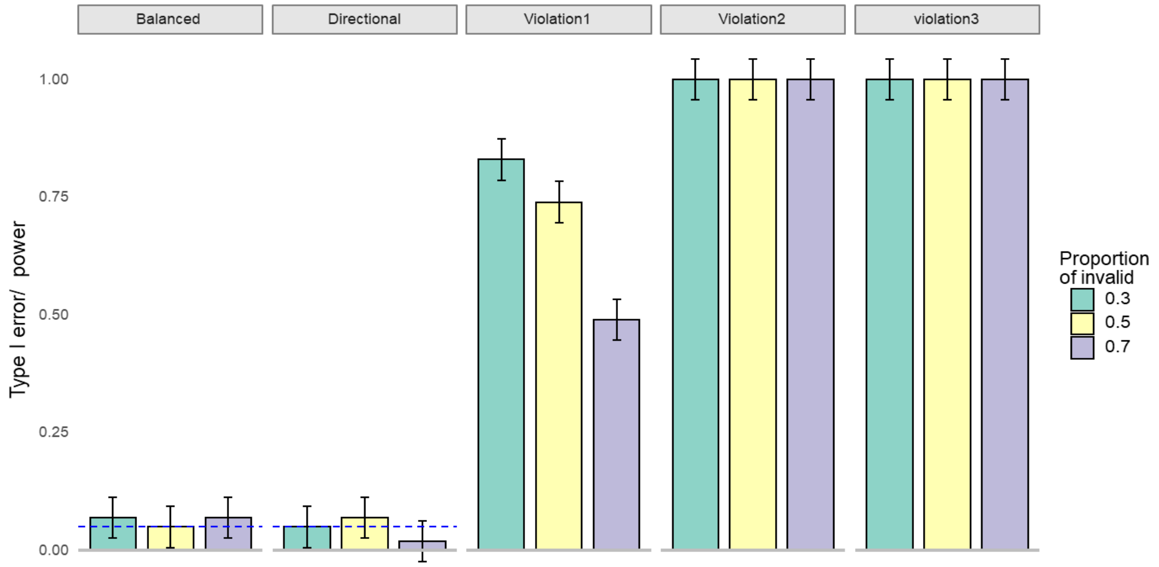


**Supplementary Figure 12: Type I error and Power of the InSIDE assumption violation test.** The first two columns (i.e., balance and directional) are simulated under InSIDE assumption holds (i.e., type I error), while the last three columns are the simulated under violation different levels ('i.e., power). The blue dash lines are expected values for type I error. The sample size is 50,000. The number of SNPs is 100, all of which were initially provided to the methods.

Supplementary table 1: Novel directional and InSIDE tests for real data.

| traits | | p-value | |
|---|---|---|---|
| exposure | Outcome | directional test | InSIDE test |
| BMI | MetS | 7.28E-02 | 2.89E-01 |
| BMI | MetS score | 2.89E-01 | 1.34E-14 |
| LDL | CAD | 2.02E-02 | 9.96E-01 |
| HDL | CAD | 7.57E-01 | 9.97E-01 |
| SBP | CAD | 8.43E-01 | 1.00E+00 |
| DBP | CAD | 9.16E-01 | 9.60E-01 |
